# A multi-step analysis and co-produced principles to support Equitable Partnership with Liverpool School of Tropical Medicine, 125 years on

**DOI:** 10.1101/2023.06.01.23290827

**Authors:** Robinson Karuga, Rosie Steege, Shahreen Chowdhury, Bertie Squire, Sally Theobald, Lilian Otiso

## Abstract

Transboundary health partnerships are shaped by global inequities. Voices from “global South” research partners are critical to understand and redress power asymmetries in research partnerships. We undertook research with Liverpool School of Tropical Medicine (LSTM) partners to inform LSTM’s equitable partnership strategy and co-develop principles for equitable partnerships.

We applied mixed methods and participatory approaches. An online survey (n=21) was conducted with transboundary partners on fairness of opportunity, fair process, and fair sharing of benefits in partnership with LSTM-Liverpool. We triangulated the survey with key informant interviews (n=12). Qualitative narratives were coded and analysed using the thematic framework approach. These findings were presented in a participatory workshop with transboundary partners to co-develop principles for equitable partnership, which were then refined and validated.

Transboundary partners identified being involved in agenda setting from the outset, shaping the design of research projects and theories of change as mechanisms to support fair opportunity however, funding mechanisms that shape power structures was reported as limiting fair opportunities. Fair process was supported by multi-directional, long-term collaborations with opportunities for capacity strengthening. Participants raised concerns about funder requirements and outdated language in contracting process that hindered equity. Fair benefit sharing was facilitated by early discussions on authorship to promote equity and policy influence. Funding also influenced the ability to travel and network, important for benefit sharing and fair opportunity. High paywalls limit sharing of research findings and access to research findings for many “global-South” partners.

The co-developed principles are part of ongoing reflections and dialogue to improve and undo harmful power structures that perpetuate coloniality within global health. While this process was conducted with LSTM-Liverpool partners, the principles to strengthen equity are applicable to other institutions engaged in transboundary research partnerships and relevant for funders.

## Introduction

### How colonisation has shaped research partnerships in Global Health

Transboundary partnerships in global health are inherently complex and shaped by global inequities. These inequities are often reflected within research partnership due to the imbalance of economic and academic resources, which disproportionately advantage those working in higher income settings (1–4). Inequities in partnerships are rooted in socio-political and historical structures of inequity and colonialism, but are maintained by the current ‘status quo’ in global health research structures and funding steams. Global Health funding predominantly comes from high income countries (HIC) and partners in the global north are often the lead grant recipients (3). As such, Northern partners may hold power over research conducted in, and for, Low- and middle-income countries (LMICs) - the global South. Literature shows how the allocation of funding can contribute to unequal decision-making and division of labour (5–7). Northern researchers therefore often have more opportunities to shape research and provide intellectual inputs, and at worst appropriate local data. Khan *et al*. write that global health organisations may “*perpetuate the power imbalances they claim to rectify through colonial and extractive attitudes, and policies and practices that concentrate resources, expertise, data and branding within high-income country (HIC) institutions.”* (8)(p.1).

Global health is burdened with the weight of its colonial roots and recognising that language has the power to harm and reinforce systemic injustice and perpetuate a false hierarchy, semantics hold weight and require critical reflection and adaptability (9, 10). In this paper, we mainly use the term transboundary research, but also global North and global South, when we wish to discuss the location of researchers. We discuss our rationale and choice of language further in our findings. We also acknowledge that “Tropical Medicine” is another homogenizing and problematic term (11).

### Liverpool School of Tropical Medicine (LSTM): Looking backwards and looking forward

2023 marks 125 years of Liverpool School of Tropical Medicine (LSTM), the oldest school of Tropical Medicine in the world created after an appeal from Joseph Chamberlain, Secretary of State for the colonies to address “mortality and morbidity arising from endemic ‘tropical diseases’” (12). The intention was both to advance teaching and research on tropical medicine in Britain, and arguably, to facilitate political and economic exploitation by Western powers and expand European colonial empires (12, 13) (14) (15) (16). As argued by Burgess (2022), we need research, and researchers, to acknowledge how the past governs and shapes our efforts to change health (17). Thus, this landmark calls for a critical and collective reflection of an organisation rooted in colonial and tropical medicine. Staff in institutions such as LSTM- Liverpool, must critically reflect on, and acknowledge how its history shapes inequalities today, to disrupt current knowledge asymmetries, through sharing relative power, examining working practices and learning about what matters to partners in working relationships. LSTM - Liverpool is currently engaged with transboundary partners through: established research and education collaborations, involvement in organisation development, having staff embedded in overseas organisations and overseas LSTM offices and is committed to fostering equitable global partnerships in research, education and organisational development.

### The future for research partnerships in global health

There is now no debating the need for change within the discipline of global health (17). There are calls to level the research playing field for research and promote epistemic diversity by ‘aligning the positionality and the gaze of global health funding models’, enhancing representation of researchers from LMIC settings through diverse research teams and overhauling editorial structures that restrict access to knowledge and privilege high-impact journals in the global North (3, 18, 19). Change involves the dismantling of deeply entrenched power structures and is no small task as power manifests itself in a myriad of intersecting ways. As Burgess and Weick write, social change can happen by ‘small wins’ - tangible acts that add up to more than the sum of their parts (17, 20). One ’small win’ is the promotion of equitable partnerships; when transboundary partnerships prioritise research needs from the global South, and embed research capacity development within partnership aims, those with fewer resources benefit from such collaboration (21). Despite the recent explosion in guidelines for partnerships that speak to efforts being made at an operational level to work towards equity, Southern authors perspectives are often missing (2, 22) 21, 22). Voller *et al.* note that Southern stakeholders “*continue to be under-represented in guideline development”* and found only two guidelines out of 22 identified documents were developed predominantly or exclusively drawing on Southern stakeholders as participants (23, 24). This perpetuates colonial structures of knowledge generation and demands attention. As Audre Lorde famously wrote *‘the master’s tools will never dismantle the master’s house’* (25) - a powerful metaphor which has been recently applied to decolonising global health (26).

Aligned with efforts to decolonise and reflect on the global health climate we aimed to co-create institutional guidance with transboundary partners to promote mutual learning, inform practice of equitable partnerships and co-develop a set of principles for equitable partnerships to support accountability and promote trust. While the exercise was conducted with LSTM-Liverpool’s partners to hear their perspectives and co-create principles to strengthen equity they are also applicable for other institutions, research partnerships, and development partners/funders working towards equitable partnerships.

## Methods

We used mixed methods and participatory research approaches to co-create knowledge, principles and encourage critical reflection and dialogue on our findings throughout the research process. Data collection took place from October 2021 to November 2022 and engaged individual respondents from 20 partner organisations, across 15 different country contexts in Africa, Asia and the Middle East. We employed several methods in succession (Figure 1). These individual methods were designed to feed into one another and helped to triangulate findings across multiple methods and perspectives, over a period of six months. These steps are described in more detail below.

**Figure 1:**
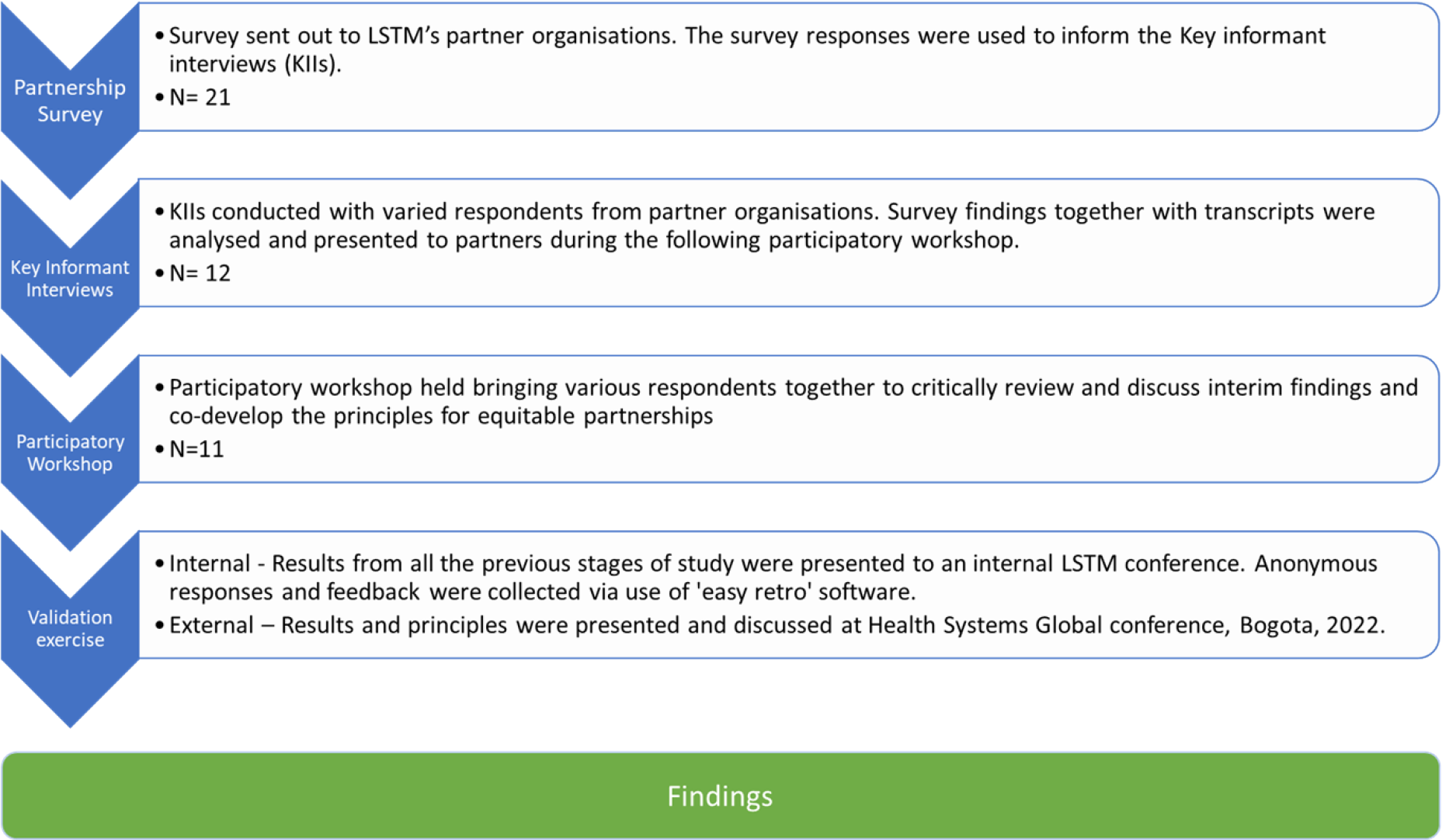
Equitable Partnerships study process – demonstrates how each consecutive stage of process informed the following stage

### Step one

We developed an online survey questionnaire for use with transboundary partner organisations to seek anonymous responses. The aim was to elicit an understanding of what LSTM and partners do well and where LSTM and partners have gaps in equitable engagement. The survey was based on, and adapted, questions from the three domains listed in the Research Fairness Initiative’s (RFI) reporting guide (27). The guide seeks to improve the fairness, efficiency and impact of research collaborations globally so it was an appropriate and standardised starting point. The domains are 1.

Fairness of opportunity (before research), 2. Fair process (during research) and 3. Fair Sharing of Benefits, Costs & Outcomes (after research) (27). The questionnaire can be found in supplementary file 1 and includes Likert scale questions and free text.

*Recruitment:* Only organisations which have a pre-existing partnership with LSTM were approached. A table of partner organisations was collaboratively developed across LSTM to ensure we captured a range of partnerships – from multiple contexts, areas of focus, type of partnership (e.g teaching or research) and length of engagement (long term engagement with LSTM vs short term /newer partnerships).

An email introducing the study was initially sent from LSTM- Liverpool’s Dean of Clinical Sciences and International Public Health (BS) to senior leads at organisations that partner with LSTM (from a range of geographical contexts) to gain their approval to participate in the study. If they wished to participate they were invited to respond directly to RK or RS confirming approval to participate. RK and RS then sent a survey link that they were asked to cascade within their organisation to capture diversity. We had 21 responses, from eight organisations working across 11 country contexts. These responses then informed the discussion guides for the Key informant interviews (KIIs).

### Step two

Qualitative interviews were undertaken with purposively sampled key informants (n=12), who were based in Africa and Asia, to gain diversity in organisations (long terms vs. short term engagement with LSTM), gender, level of seniority and a range of geographical contexts. Interview guides were co-developed by researchers from LVCT Health (Kenya) and LSTM (UK) and focused on several areas relevant to partnerships such as research agenda setting, impact of the partnership on global and local policies, funding structures, capacity development initiatives and language concepts. Interviews triangulated findings from the survey and explored issues in more depth to understand which ways of working currently serve to amplify partner’s perspectives and support equitable engagement and whether partnerships with LSTM impact on the broader contexts we are working in.

*Recruitment:* Survey respondents were asked to indicate if they/or another member of their team would like to participate in a KII and we recruited some participants this way. Additionally, emails were sent out to gauge interest to participate in interview from those who had not participated in the survey but had indicated willingness to participate in the study. RS and RK conducted the interviews virtually via Microsoft Teams or Zoom and informed consent was obtained. While authors had access to identifiable information about participants due to the participatory nature of the study, all data was anonymized after data collection.

### Step three

We held a collaborative workshop in March 2022 with LSTM’s partners (n=11) to co-develop a set of principles for LSTM partnership. Invitations to an online co-development workshop were sent to all organisations who agreed to partake in the study, this meant we had some attendance from people who were interviewed or participated in the survey but largely attendance was from people who we had not previously spoken to providing diversity of respondents. The workshop allowed us to disseminate interim study findings and begin the process of defining a set of principles equitable engagement with transboundary partners. This was facilitated by external partners at LVCT Health, Kenya (LO, RK) and staff who were not within senior LSTM management (RS, SC). Senior LSTM colleagues (SBS, ST) joined at end of the session to affirm their commitment to the process and give thanks for partners’ participation in the process.

*Recruitment:* A date, time and meeting invite was advertised via email to all organisations who had indicated willingness to participate in the study.

### Step four – validation exercises

An internal validation session at LSTM staff hybrid conference (n=173) (included staff based in Liverpool and Kenya, Malawi, Nepal, Uganda and ZImbabwe) was held to disseminate findings and draft principles. We collated inputs and comments via easyretro (https://easyretro.io/) to inform principles and associated actions. We received 114 inputs which were themed by two different analysts (SC and RS) and reviewed alongside the empirical data to inform our analysis, but not included as empirical data.. We also undertook an external validation exercise via a ‘world café’ approach at Health Systems Research (HSR) Conference, Bogota November 2022 (approximately 40 participants). This provided an opportunity to present the co-developed principles to a wider audience to discuss and refine them. There was general consensus that the principles were appropriate but questions were raised about accountability for principles, including processes and resources for monitoring and learning, which we set out in the discussion. Finally, the amended co-developed principles were shared with all study participants for final comments.

#### Data Analysis

Simple descriptive statistics were used to identify key trends in the survey data, and free text questions from the survey were coded in NVivo version 12 (QSR International, Australia). Interviews were transcribed verbatim and collated with free text responses from the survey. Data collected was validated, organised and summarised according to themes. We used an inductive framework approach, through sorting data with coding and followed by exploring and explaining links between codes and themes (28) facilitated by NVivo 12. The participatory workshop with transboundary partners (n=11) served as participant checking on the analysis as well as an opportunity for co-creation of the principles. The LSTM Staff conference responses and easy retro inputs were reviewed alongside the analytical process as an internal validation component that shaped our final principles. The final principles were also externally validated by diverse health systems researchers and actors at HSR, 2022. Three researchers with varying positionalities came together to validate coding structures and summarise themes (RS, SC, RK).

#### Ethical considerations

This study received approval from the Research Ethics Committee at LSTM (21–060) in September 2021. Participant information sheets were provided, and written consent was obtained for all workshop participants and for those who provided written responses.

#### Positionality, power and engagement

The survey allowed for anonymous responses to be gathered so participants could be critical. The positionality of interviewers was also important to mitigate against this – interviews were either conducted by LVCT Health staff (RK) – a Kenyan based organisation external to LSTM, or a previous LSTM staff member (RS) who had not been part of senior management at LSTM and was independent from LSTM at the time of interviews; both interviewers felt that they were able to build rapport and have open and critical conversations. Further, a general positive perception respondents held of working with LSTM may have facilitated critical discussion of the power imbalances within global health research. We felt participants were comfortable to discuss the uncomfortable, which was essential to have meaningful dialogue on this.

## Results

We gathered responses from participants working across 20 different organisations and 15 different country contexts across the stages of the research process (table 1).

**Table 1:** respondent characteristics across the three stages of the research process * Given the survey was anonymous we do not know how much overlap there was between survey participants, key informants and workshop participants therefore we are providing a range of total participants.

| Method | Survey (n=21) | KIIs (n=12) | Participatory Workshop (n=11) |
| --- | --- | --- | --- |
| Organisations | N=8 | N=12 | N=10 |
| Gender | Man (n=13)<br>Woman (n=7)<br>Other identity (n=1) | Man (n=9)<br>Woman (n=3)<br>Other identity (n=0) | Man (n=8)<br>Woman (n=3)<br>Other identity (n=0) |
| Nationality | North Africa (n=1)<br>East Africa (n=6)<br>West Africa (n=2)<br>Southern Africa (n=1)<br>South Asia (n=1)<br>Middle East (n=0)<br>Europe (n=1)<br>Dual (n=1) | North Africa (n=0)<br>East Africa (n=5)<br>West Africa (n=2)<br>Southern Africa (n=2)<br>South Asia (n=3)<br>Middle East (n=0)<br>Europe (n=0)<br>Dual (n=0) | North Africa (n=1)<br>East Africa (n=4)<br>West Africa (n=3)<br>Southern Africa (n=1)<br>South Asia (n=0)<br>Middle East (n=1)<br>Europe (n=0)<br>Dual (n=0) |
| Organisations' involvement with LSTM | 2-5 years (n=4)<br>5-10 years (n=11)<br>10-20 years (n=2)<br>>20 years (n=1)<br>Don't know (n=2) | 2-5 years (n=1)<br>5-10 years (n=5)<br>10-20 years (n=5)<br>>20 years | 2-5 years (n=1)<br>5-10 years (n=4)<br>10-20 years (n=4)<br>>20 years (n=2) |
| Total | N=22-41* |  |  |

Results are presented first by findings on language, then by thematic areas mapping to the domains from the RFI – fair opportunity (before research), fair process (during research) and fair benefit sharing (after research). We end with co-created principles and a discussion of how these were shaped. Under each theme, findings are integrated and triangulated across the survey, interviews and participatory workshop, including recommendations from partners where applicable.

### Language Terminology

Overall, while there was a preference for the term ‘global south’ there was no clear consensus. One participant emphasised that the language was not important as the different UN agencies and multilaterals all have different mechanisms by which they categorise and label countries, so it is difficult to achieve consensus on categorisation even within terms. Categorising countries by their geographical location, or economic status in either way was viewed to be inaccurate, but inaccurate geography was viewed more favourably. The term majority world was also offered as an alternative in the workshop.

> *‘Again, I think the LMICs for me is not is not palatable… it’s just a label…it’s not realistic. [We] might be a poor nation, but now we have got millionaires…’ (*KII – senior, man, Southern Africa)

### Fair opportunity - Funding streams

While the majority of survey respondents (17 out of 21) agreed that the partnership with LSTM attempts to ensure that research funder demands do not cause unfairness in partnership, three respondents disagreed. This may be reflective of the funding structures that give LSTM more power within the partnership that emerged in KIIs. Partners reported that most projects in the partnerships were funded from sources within the UK. This was described as shaping the power dynamics in the partnership, mainly attributed to LSTM’s interaction with funders, and should be an area of reflection:

> *‘[LSTM] are the ones who deal with the donors, because that’s just how our funding is arranged. So, there will always be more power for the people at LSTM, in terms of taking charge of the project… And all the projects that we’ve done, they have always been prime’ (*KII – Senior, Man, Southern Africa)

Budget allocations were described as being discussed collaboratively and transparently. Partners reported LSTM supported with financial management capacity, technical and liaison support when engaging with funders. All partners were affected by the UK’s 2021 Overseas Development Assistance (ODA) funding cuts following COVID-19. Partners reported having open and transparent discussion on how to work with limited funds after the 2021 funding cuts and reported positive outcomes of LSTM Principal Investigators (PIs) negotiating down the size of the cuts. Unsurprisingly, funding structures that rely on Northern funded research were described as stifling global equity; domestic research budgets were described as being critical to equity:

> *‘…So, in terms of decision making, the global south has been included in decision making in many ways. But we might be disadvantaged, or we might not reach the absolute end of equity, if we are not financially independent. So, I think that if we really want to maintain independence, then countries in the Global South should begin to think of how they can generate research funding from within… So, I think in terms of resource capacity, that is one area that might likely stifle global equity.’* (KII- Senior, Man, East Africa)

It was suggested that LSTM can play a role in facilitating dialogue between UK funders and LMIC partners to foster transparency and build the capacities of both funders to understand contextual realities in the global south, and of partners in understanding funding mechanisms. Further, through our participatory workshop, it was highlighted that the space to bring together different partners who do not normally come together was valuable and something LSTM-Liverpool could continue to facilitate to support fair opportunity.

### Fair opportunity - Agenda setting

Agenda setting between LSTM-Liverpool and partners was often described as being defined mutually and often a participatory process, particularly when physically together. Proposal writing workshops and early engagement with partners was seen to help implementation and sustainability. Partners mentioned being involved early in the partnership(s) and able to input into the aims, objectives, methods for the partnership proposal and co-develop the Theory of Change within contextual realities.

> LSTM is really good at ensuring that its partners are engaged as early possible on any opportunity that arises. This has led to not only smooth implementation of the project (s), but implementing very successful projects and partnerships where each partner is satisfied and even wishing for more. (Survey respondent, man, Southern Africa)

Agenda setting was also mentioned as being defined by the funding call and the global funding architecture, which is predicated on global power dynamics. Ensuring this power dynamic is then not replicated within the partnership itself was highlighted by one participant.

> ‘Somehow our partnership is also influenced by the global agenda setting process… our partners in higher income countries who dominate in the selling process. However, I’m not blaming, but that has some kind of synergies or linkages between the partnership of two institutions like [organisation] and LSTM team. However, understanding of the global agenda setting process, not repeating the weaknesses of that process, while agenda setting between two partners is very, very important… open communication really that’s the first thing …and not dominating the discussion is very important.’ (KII-Senior, Man, Southern Asia)

> Uh, and I think that’s driven to a large extent by the fact that high income countries are putting in more money and therefore they are answerable to their parliaments and not [our] parliament. I don’t think we will get into a situation where that is completely balanced out. I’m not even sure that it needs to be completely balanced out.’ (KII- Senior, Man, East Africa)

Agenda setting in response to local needs was reported to be a balance of country priorities and funder influences. It was noted that the UK funding landscape is shifting to allow a broader focus which gives more space to align with country priorities. Long term engagement and familiarity with contexts means that partners are able to respond to local needs in priority setting. One participant who had a longer term engagement with LSTM also noted that the response to local priorities had been given more weight in recent years, whereas historically LSTM’s priorities had been dominant. Despite funding streams generally flowing from North to South, there were examples where Southern organisations were the consortium lead and funding recipient.

> *‘We do the agenda jointly. I mean, [consortium name] is leading the research uptake, the expectation from the team is that Africa leads that program, leads the consortium, but, leading doesn’t mean that you determining everything, you basically co-create within the consortium. But we lead those components, and so the agenda is set by the consortium itself.’* (KII-Senior, man, East Africa)

Space for innovation within protocol and grant writing was described as a current gap as many partnerships are focused on executing research via traditional methods. This raises the question of what spaces can be created for co-production and co-creation through prioritising local knowledge.

### Fair process - Collaboration with LSTM - Liverpool

Collaboration between partners and LSTM-Liverpool took different forms, namely research, capacity strengthening, teaching, research uptake, advocacy and publication. Length of engagement and physical proximity were noted to influence the collaboration. One respondent noted the benefit of LSTM-Liverpool having a physical in-country presence when collaborating, as opposed to only attending for meetings. Physical separation was reported to negatively impact collaboration as institutions are perceived to not “*walk the talk”* (KII- Senior, man, Southern Africa). The importance of being physically together, through university faculty exchanges, was also stated as a way to strengthen the partnership, but it was emphasised that this should be a two-way process.

The personality of PIs from LSTM-Liverpool were also influential on the nature of the collaboration. Some PIs were mentioned for their excellence and commitment to equity in partnerships, which influenced a valued, personal relationship with LSTM projects. How much this is reflective of a culture of fairness within LSTM-Liverpool is unknown. Multi-directionality was also stated to be an important factor in the nature of the collaboration.

> What I like about [consortium] what’s been good, we had [name of PIs] and [PI name]’s approach is very different… So you know, in some cases, …….there is engagement, you do great research, but there isn’t that feeling of you’re part of a community. But with [consortium] I feel like we’re part of a community …(KII - senior, woman, South Asia)

> Actually, we have a rotating chair, I think way different partners take charge in chairing the management meeting. This shows that there are efforts to make sure that partners especially in the south involvement in the project. (KII-ECR, man East Africa)

General decision making within partnerships was described by the majority of participants as fair and respectful. Nevertheless, two survey respondents felt power dynamics that influence decision making, are not always explicitly discussed and roles are not always clearly defined. Developing organograms and advocating for rotating chairs in meetings emerged from KIIs as valuable to understand who will lead what and share power at the outset.

It was also noted that LSTM-Liverpool has played a role in establishing transboundary organisations. While LSTM-Liverpool’s supportive role was acknowledged, the analogy of LSTM-Liverpool as a parent and the partner’s organisation as the child was used to describe the relationship, suggesting a paternalistic collaboration with links to a colonial past. The analogy was also used to state the need for independence, suggesting how although partnerships, capacities and relationships evolve through time, power dynamics can be more entrenched.

> “I think [of LSTM] as our parent, we take ourselves as, as the brainchild of the LSTM… over the years, we’ve seen LSTM supporting staff from [partner name] with education opportunities, master’s degrees, PhDs and I happen to be one of those are beneficiaries of this partnership… So they’ve seen us through thick and thin. And that’s why I use the term - like a baby, LSTM as a parent…What LSTM is trying to achieve should actually also be seen in practice…you know, there reaches a point whereby a child is grown and wants to walk alone. And I know that’s very hard for the parent to let go. But sometimes you do have to let go and just watch from afar. So maybe that’s where LSTM and [organization] are.” (KII- senior, man, Southern Africa).

To support equity in fair process a suggestion was to leverage virtual mediums for knowledge exchange and networking to expand and develop collaborations, as these can enhance equity as costly travel and visa complications are avoided.

### Fair benefit sharing - Capacity strengthening

Capacity strengthening was seen as a strength of LSTM-Liverpool’s engagement. It occurred intentionally and evolved naturally by nature of the partnership. There was value placed on data sharing and post research agenda, documentation, new methods and communication of research. Project length was described as an important factor in influencing the success of capacity strengthening initiatives, with longer projects e.g. 5 years, allowing space for individual capacities to be enhanced.

> *‘…projects that are two years you just getting into the momentum. And then it’s like see you later, goodbye, wrap up… But is that when you have a longer term project, I mean, colleagues come and go, but the few that you can see them grow in terms of their skills. And I think that’s very good.’* (KII - senior, woman, South Asia)

Capacity strengthening was mostly focused on an individual level particularly from LMICs, with MScs, PhDs and post-docs being highlighted as particularly beneficial for mid-career researchers. Careful attention however needs to be paid to the distribution of academic opportunities within consortium partnerships to support equity:

> Training needs to be equally considered in participating countries for example all PhD opportunities went to two countries while we had more several participating countries. A selection criteria need to be purposeful as every country needs to strengthen research capacity (Survey respondent, East Africa)

It was also noted that projects with a transparent and horizontal hierarchy structure have benefits to individual capacity strengthening – particularly for early and mid-career researchers. Individual capacity strengthening has also evolved from community engagement - ‘unlearning academia’ - these projects were reported to develop skills in blog writing, photo narrative writing and podcasts.

> Capacity strengthening has really been at its best since we started collaborating with LSTM. We have our capacity building in different areas relating to research, to communication, to research methods, … it has been a wide range of opportunities for capacity building and it has really enhanced our work, even in different projects that are not related…’ (KII- ECR, man, West Africa)

Funding for institutional capacity building was identified as an unmet need but was reported as critical for establishing credibility for organisations. One positive example was mentioned whereby staff were sent to do secondments at LSTM-Liverpool and returned to build up institutional capacity at home. Critically however, it was noted that this should be multi-directional.

While some participants mentioned improvements in budgeting and financial management as a result of the partnership, many partners mentioned the need for capacity strengthening of financial management systems, research and management capacity as well as publication for high impact journals. Survey results suggested that there are gaps in the provision of resources for capacity strengthening of project staff and it was also acknowledged that administrative and project management staff are often overlooked. This is a critical area for consideration as one participant emphasized the need to acknowledge transboundary partner institutional and contextual limitations which may impede on their opportunities to lead partnerships and deliver to specific templates for reporting.

> *‘We are an organization established and evolving and struggling in developing world with lots of limitations … resource limitation, limitation in terms of capacity, limitation in terms of the institutional component where you require investing quite a lot in terms of building institution, not necessarily that is up to the standard of the LSTM institution But using a standard global templates that are the template for all one size fit for all, while setting the standards from the LSTM side… that presents limitation to organisations like us, we are not as big as LSTM, but big does not mean that it should dominate the standards…I think it’s important that there is a rigid trust built in both end in terms of managing a project under this partisan framework. And understandably, it takes time to build that trust… in order to develop trust both end by knowing the institutional practice….’* (KII Senior, Man, South Asia)

Recommendations included completing a competency assessment to establish capacity strengthening needs in research partnerships and to onboard all partners with established and clear templates for planning, budgeting and reporting. One participant suggested that LSTM-Liverpool could support partners with access to academic and online training courses currently available only to LSTM-Liverpool staff and students e.g. in safeguarding, and access to libraries. It was also emphasised a focus on strengthening capacity of partners to innovate and think outside of the traditional research process would be beneficial.

*So I think it’s the capacity to understand where innovations can be…there are small things such as theory of change, etc. These are things which are not normal in our traditional medical research. So it’s something we’re also taking on board, it’s more of a global health issue, public health. So these are things we’re learning as we move along. So.., the UK may be way ahead of us, but we’re learning so even when we write proposals, we always gain knowledge. They understand how things work on the ground, what the problems are, but we also gain that understanding of how to write certain proposals*. (KII – senior, woman, East Africa)

### Fair benefit sharing - *Impacts and outputs*

Research uptake at the local and national level is reported to be led by partners, whereas LSTM- Liverpool was reported to facilitate at global level - including publications. The focus on LSTM’s impact at the global level reinforces broader power relations in knowledge generation within global health and does need to be re-evaluated in order to support equitable partnerships. Nonetheless, impacts of projects were described as having a major contribution to the policy dialogue and partners’ strong relations with the Ministries of Health were stated to be valuable in raising visibility through international consortia. One partner also stated their existing strong relationships with policy makers at national and local level should not be undervalued in creating policy change. Another spoke to how the strengths of each institution come together for impact:

> *‘our strength lies with actually the understanding of the context…In terms of the problems and the gaps and how to be able to address them…You have good ideas, but good ideas if it’s not funded, it will just remain good ideas* (KII - senior, man, West Africa)

Authorship with LSTM-Liverpool was described as fair and equitable and commendable. Good practice involved early discussions on equitable authorship, a commitment to follow journal protocols and guidelines and utilising opportunities for joint first and last authorship positions. It was noted that the publication process is usually led by LSTM-Liverpool and northern partners, and that this is also shaped by funding, limiting equitable access to publishing. “*To be honest, I can’t afford $5000 to publish in maybe in BMJ or so*” (KII -Senior, man, West Africa). It was felt generally, academics put more focus on outputs and publications while non-academic partners understand the local context and influence policy in different ways other than publication, but this is not always considered as being of equal value.

A critical area of concern however, with regards to impact and uptake is insensitive and outdated legal language in contracts with LSTM-Liverpool – which often state that data is owned by LSTM- Liverpool. This was described as promoting lack of ownership, and limits opportunities for Southern led authorship and demands urgent attention across Northern institutions.

> *‘Legal language in contract that data is owned by LSTM, data should be owned by researchers with right to publish. Promotes lack of ownership.’* Survey respondent, Woman, East Africa

This highlights the need for data sharing agreements to be put in place which give LMIC partners the power and rights to their own data and grant LSTM access to support in dissemination and power sharing. Further suggestions to increase equity in fair benefit sharing was for LSTM-Liverpool to leverage their position to increase power of Southern partners by introducing and nominating partners to be involved in international forums such as technical working groups or conferences, or by promoting LMIC partners in publications. Additionally, considering opportunities for publishing other outputs such as blogs and policy briefs alongside traditional journal articles.

### Co-developed principles for equitable partnership

The values that survey and interview participants identified as critical to equitable partnerships are depicted in Fig.2. The majority of respondents agreed that their partnerships with LSTM-Liverpool matched these values. LSTM-Liverpool was regarded as doing well in equitable partnerships through early collaboration and engagement, support for staff, honest and transparent processes and a focus on co-leadership.

**Figure 2.**
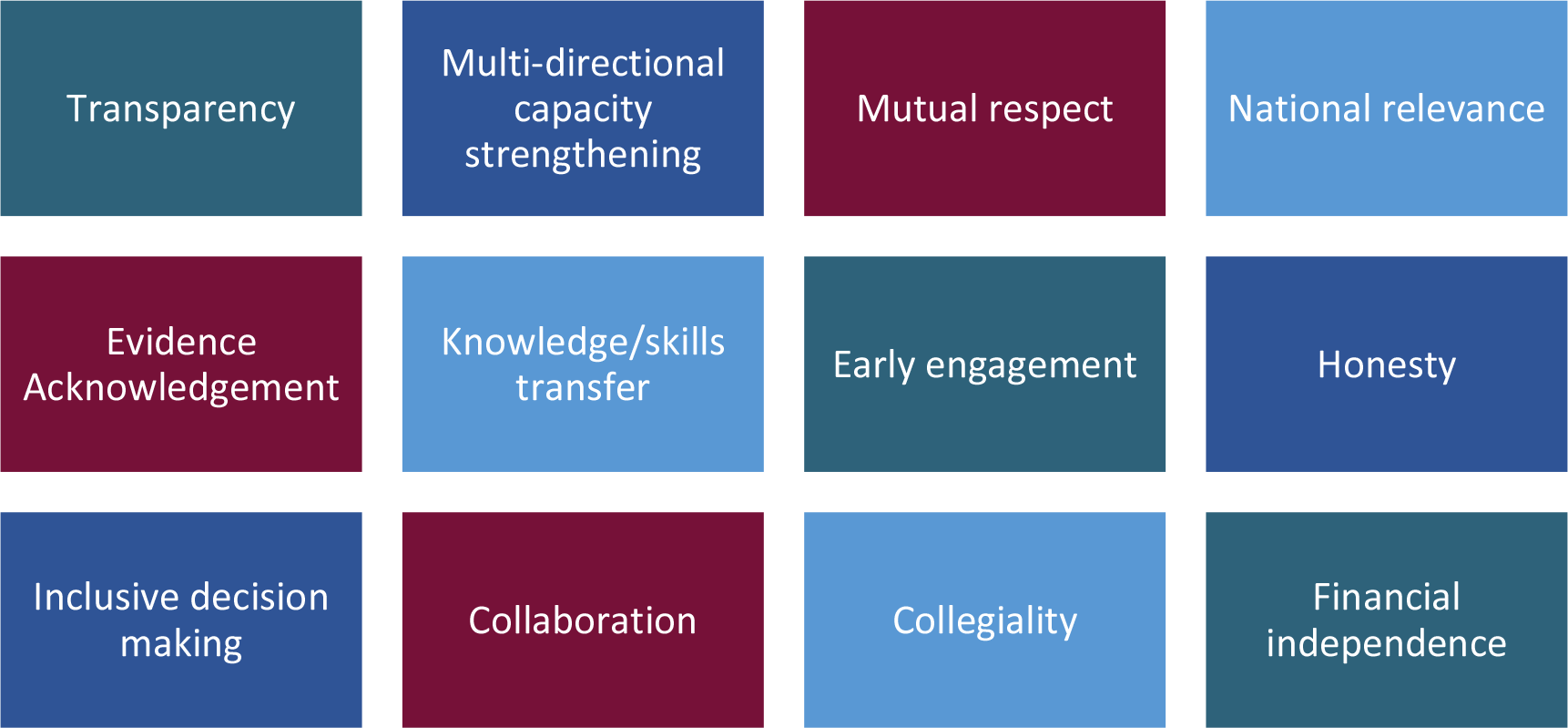
Synthesis of values of equitable partnerships from KIIs and survey findings, presented in participatory workshop with partners, which informed the draft principles.

These values were then shaped into draft principles by (RS, RK, SC) to present in the participatory workshop for discussion and input. Partners within the workshop further edited and refined the draft principles. Key edits pertained to capacity strengthening being multi-directional (principle 6), and the inclusion of values of the Global Code of Conduct (principle 7). These principles were then shared again via email and further refined. Final principles are shown in Box 1.

Many partners suggested that equity could be evaluated against agreed parameters, through follow up interviews over time as well recorded documents, regular feedback and record project evaluation from both sides and that accountability mechanisms for voicing concerns exist, through open communication (particularly when partnerships have been built over the years); regular management meetings, safeguarding protocols and working groups were described as forums where partners felt they could voice concerns formally or informally. It is important to note however, that this was stated by respondents who work at senior levels.

**Box 1:**
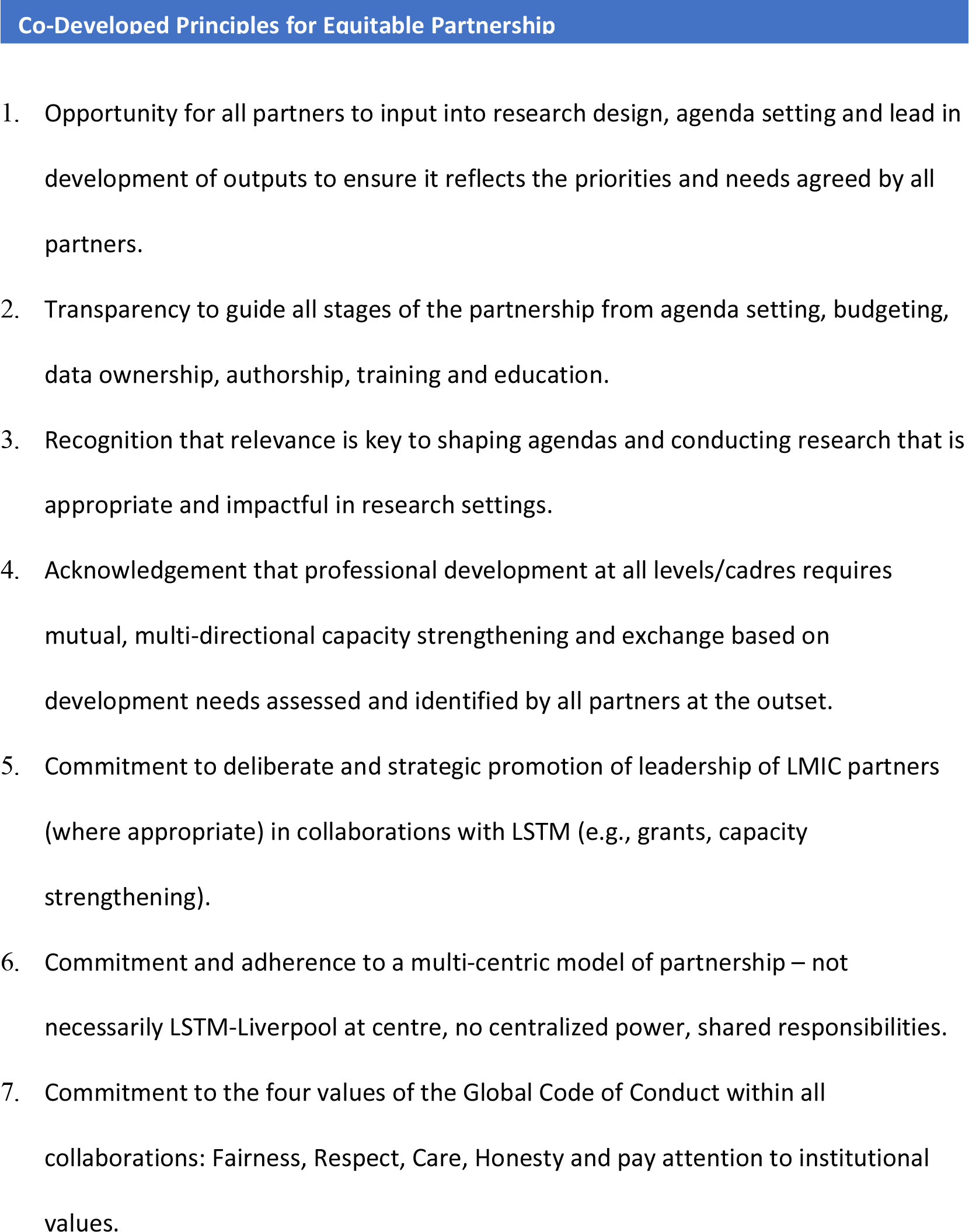
The co-developed principles for equitable partnership

## DISCUSSION

This study is the first-time LSTM-Liverpool’s partners have been actively engaged to co-develop institutional principles to support meaningful and equitable engagement. It seeks to centre Southern perspectives in progressing equitable partnerships and dismantling the ‘master’s house’ of global health.

### Our findings

We found that research partnerships with LSTM were overall, viewed positively, and equitably. With regards to fair opportunity there were positive accounts of partners being involved in agenda setting from the outset able to shape and input into the design of research projects and adapt theories of change to their contexts. Fair process revealed that the collaborations were viewed as multi- directional, positive and personal. Causes for concern around funder requirements creating unfairness within the partnership and outdated language in contracting and partnerships not supporting equity were also raised in the anonymous survey responses and require further exploration. There was an emphasis on capacity strengthening in LSTM-Liverpool’s partnerships that was valued by partners, though it was highlighted this needs to be extended to research support staff. Finally, fair benefit sharing revealed equitable processes around authorship supported by upfront discussions and policy influence. The interlinkages between the three domains, and how they are situated within broader political economy of research partnerships also clearly emerged in our findings. For example, funding structures underpin power structures that dictate fair opportunities, both in research grants and shaping the research agenda. Funding also influenced ability to travel, network and share findings. This was important for both fair benefit sharing but also fair opportunity as it was reported many collaborations with LSTM-Liverpool started through networking at conferences internationally. Funding also influenced fair benefit sharing as high paywalls limit both sharing of research findings and access to research findings for many Southern institutions.

Additionally, findings from some KIIs had clear links to a colonial past, for example the language of LSTM being the parent, speaks to paternalistic collaborations which Okeke has dubbed ‘the little brother effect’ and requires LSTM-Liverpool staff, as well as staff in other northern institutions, to be critically reflexive on their well-meaning role as ‘older brother’ (29). Colonial assumptions about knowledge superiority were also discussed where partners described how capacity strengthening opportunities usually flowed from north to south. Southern staff would go to do secondments at LSTM-Liverpool and other northern institutions, but it was less expected that northern staff would come to learn at Southern institutions. This highlights the importance of the multi-directional capacity strengthening and exchange initiatives for equitable partnerships, which was stressed and is reflected in co-constructed principles (for example, principles 4-6) (30). There was also debate around accepted terminology for geographical and political areas. What is clear however, is that efforts to homogenise large swathes of geographies into clear and widely accepted terminologies fails to capture the nuances and the varying facets that characterise privilege and disadvantage in those settings. Instead, we should embrace ‘pluriversality’ and stop trying to reach consensus, which it has been argued is required for decolonization (31). It is worth bearing in mind however, that language is not the sole issue, rather we need to think critically about the multiple power relationships that language represents and tailoring language may be appropriate (32) (10). The process of engaging in reflection, dialogue and debate is one of the ‘small wins’ we can make on the journey towards changing existing power dynamics (9, 20).

### Principles

We co-developed seven actionable principles to guide equitable research collaborations and validated these with different audiences (See Box 1). The process taken to arrive at the final principles presented here took various methods and iterative consulting (figure 1). These principles reflect some of the key issues/areas that are centre stage in other definitions or approaches (for example there are some similarities in the UKCDR definition with a focus on mutual participation, mutual trust and respect, mutual benefit and equal value placed on each partners contribution at all stages of the research process). The process itself was also of value, where LSTM-Liverpool and partners took time to deliberately reflect and define the partnership, a process that should be maintained regularly. Partners also noted the value in being brought together to share ideas outside of the confines of existing research partnerships, a process that it was suggested LSTM-Liverpool continue to facilitate.

It is important to note that these principles must not be viewed as operating in a vacuum. We, as researchers need to engage with the research eco-system that shapes the partnership, creating space for mutual learning and adapt to changes in the external context (23). For example, partnerships are influenced by the changing global funding architecture; the recent climate of UK Overseas Development Aid (ODA) funding cuts amidst a global pandemic, brought unexpected budget cuts and additional challenges to transboundary research as highlighted by respondents and in recent literature on research partnerships (33). Individual and institutional action is critical but also needs to be part of a wider process of change within the broader political and funding ecosystems that shape and underpin research partnerships through funding infrastructure (donors) and knowledge dissemination avenues (journals). Indeed, Voller *et al* (2022) notes how guidelines may not always fully acknowledge the structural barriers and competing interests that get in the way of these changes being realised (2).

### What the implications of our findings – for LSTM-Liverpool and partners; and for broader research infrastructure

#### Role of funders and the future of funding

The funding structures in research, that typically flow from north to south was lamented by our respondents as inhibiting equitable partnerships more generally. This speaks to the need for political change and for the funders to engage with, and embrace, the complexity of equitable research partnerships and for funders to embrace contextually specific research to advance health systems research. This requires a shift from Eurocentric modalities of research funding to support Southern led research and encourage co-production approaches with populations in these settings, which is occurring (34). Some funders are now funding LMIC institutions directly, as well as setting up development funds to foster and enable partnership working in the grant development process (e.g. NIHR, UKCDR) which is to be applauded. However, funders need to pay critical attention to ensuring that funding provision is equitable and that by funding Southern institutions directly, they do not leave them worse off by imposing excessive rules, regulations and costs that the organisations may not be able to bear. As highlighted by respondents, institutional capacity is not always as strong as large academic institutions and as such there needs to be flexibility in reporting requirements. Funds for embedded operational capacity strengthening and ongoing work into distilling learnings on equitable partnership are also critical – these must be co-created with Southern institutions, so it has value for them and not just based on the funders needs.

As described by one of our respondents, investment into locally led research is also critical. Domestic sources of funding for research is critical to mitigate against power imbalances in global health research and should be what we strive for. The political climate of many northern countries may even force this – recent UK ODA funding cuts were a source of concern for many partners we spoke with and risked undermining relationships with UK institutions. Further, the UK and Italy were notably absent from commitments to the Global Fund in September, 2022 who missed their replenishment targets (35). Despite their absence, many Southern countries did pledge, some such as Malawi, for the first time, which speaks to a shift towards who is funding global health with implications for equity.

#### Knowledge generation and dissemination: Role of journals, conferences and beyond

Knowledge systems in research and academia need critical review. There is a growing body of work on research equity, authorship, and the role of academic journals in the context of international health research partnerships - see for example the “Consensus statement on measures to promote equitable authorship in research publication from international research partnerships” (36) . A number of journals (for example Health Policy and Planning) are now including reflexivity statements, which require author teams to critically reflect on roles and responsibilities and demonstrate inclusivity by regional location. Equity in authorship and in access to papers has also been a key theme in recent conferences (37). Hosting conferences and forums virtually, or in hybrid form, may also confer a benefit in terms of equity of participation when compared to in person meetings that may be expensive and include visa complications (passport privileges emerging as an important area for consideration). Practical suggestions to support equity from our findings were having authorship discussions up front, and making use of joint first and last author positions, as we have done in this paper. Transparency and trusting relationships between partners is therefore critical to facilitate these potentially uncomfortable discussions.

Inequities in knowledge dissemination and sharing remain, our research participants highlighted barriers in accessing resources to publish, and challenges in accessing resources which are not open access. There also inequities in who publishes: a recent review of papers reporting research trials from the global South showed from 1990-2013 papers with a first author from a LMIC increased 2.8 times while papers with a first author from high-income countries increased 11.8 (38, 39). To date, knowledge generation has favoured those in the north over the south (38). This is maintained by the so called ‘international’ reputable academic journals such as The Lancet or BMJ that are often headquartered in the global north, as raised in our external validation exercise (36). We should strive towards knowledge that is created and published within the global South as a valuable pathway for Southern led and globally disseminated knowledge. It was also highlighted how equal value should be given to understanding the context and policy space as well as academic publications, speaking to the need for structural change in how we value knowledge in global health.

#### What are the implications for LSTM and next steps?

On the nature of collaboration, the measure of an equitable partnership was sometimes measured by the merits and approach of individual PIs rather than as the institution as a whole. A key point that emerged from the respective discussions is how will the principles be rolled out and assessed? Whose responsibility is it to review adherence to the guidelines and document learning to guide future work? LSTM’s next steps involve fostering a culture among all staff (legal, financial, administration, researchers, lecturers etc) that supports and values approaches to supporting equitable partnerships. This will require relative power sharing, both in terms of securing funding but also in promotion of Southern colleagues for benefit sharing where appropriate. It may also entail ongoing review at annual staff meetings; consideration of the principles within the research proposal submission system, processes of ethics, safeguarding and research integrity and governance; an overhaul of outdated contracts (40), changes and trust in the way intellectual property and data is handled with partners, establishing multi-directional structured mentorship streams, re-evaluating academic currency and promotion criteria to value the soft skills of partnerships as well as the measurable outputs from it (5) and critically, requires the will and the drive to do it. Further, LSTM and academic institutions may wish to think about who has access to short courses and resources – whether this is limited to current students, or extended to alumni and transboundary partners. It will also be crucial for staff in LSTM-Liverpool to critically reflect on their own positionality and how to embed transformative learning in curricula to sensitise upcoming generations of staff, and build fair and transparent processes that acknowledge different people’s multiple roles and responsibilities.

The principles that were co-created cannot be another set of principles that sit in an academic archive. These have to be lived by LSTM staff and partners, and can hopefully be leveraged by partners to use in external relationships too. In order to ensure they do not simply become a ‘tick box exercise’ it is critical to have accountability mechanisms in place. During the participatory workshop it was suggested by participants that further discussion amongst groups about how partnership with LSTM would be useful, on an annual basis, possibly in a partnership council; with a convenor and a rotating chair. This possibility and funding for it will be further explored and could provide an opportunity to reflect and critically discuss the application of the principles and document learning. These principles are also intended to act as a means for accountability. They should be agreed upon at the beginning of partnerships and reviewed throughout. We also encourage other institutions to engage in a similar reflective process with partners. Acknowledging the burden on time of partners, we also welcome use of these principles for adaptation.

## Limitations of our approach

Though we began the critical process of engaging with partners to co-develop principles for equitable partnerships, there were some limitations to our approach. Firstly, we sampled from our own partners which may have introduced some bias as partners generally felt positive about the relationship. Though we sought to recruit diversity in partners, it is likely that those who had a positive view of LSTM opted into the study more so than those who felt indifferently, or negatively. Given recruitment processes were initiated through LSTM, participants may have felt they needed to respond positively about the institution, we aimed to mitigate this through interview being undertaken by LVCT Health (RK) and ex-LSTM (RS) employees. We also mostly recruited partners who had a senior role within their organisations for the KIIs and the workshop. Although we intended to engage partners from a range of career stages, the ethical approval process to conduct the study required the approval of the organization head before we could approach other researchers within the organization. Senior respondents were asked to cascade the recruitment call within their organisations but this relied on their active engagement and may have limited our respondent pool. There is a need to engage further with more junior researchers from transboundary partners to mitigate against power imbalances that may have limited their participation. We also had a fairly small sample of respondents, which again may have been limited by the strict ethics process for recruitment. The multi-method approach (Figure 1) aimed to mitigate this by capturing a range of diverse views and perspectives through time, providing a holistic picture of the nature of partnerships. The challenges of recruitment also impacted the global diversity and range of organisations that we spoke with, though our sample included 20 organisations reflective of the areas in which LSTM-Liverpool works. Many organisations who we spoke with were engaged with only a handful of PIs, so to what extent our findings reflect working partnerships across different departments within LSTM is uncertain. Finally, our research was conducted at a particular moment in time, COVID-19 had led to UK ODA funding cuts and high levels of uncertainty and precarity in relationships, with early and mid-career researchers impacted most of all. This allowed us to explore how LSTM responds to challenges but our results need to be interpreted with this context in mind. Despite these limitations, we feel that the participatory analysis, co-construction and validation processes aimed to bring diverse views and perspectives into the process and to mitigate against some of the recruitment and power issues that may have influenced the research process.

## Conclusion

Fostering equitable partnerships means we must confront uncomfortable truths within partnerships; acknowledging and discussing how funding sources, donor priorities, history and language shape existing power asymmetries. LSTM-Liverpool is in the midst of an ongoing process of deep reflection with partners. It is important to note that equitable partnerships is not something to be ‘achieved’ rather, it is the process of ongoing reflection and dialogue, seeking to do better and to dismantle harmful power structures that perpetuate coloniality within global health. These principles represent small steps on this journey and critically include the perspectives of transboundary partners. We hope these principles will be helpful to others and will be adopted by LSTM-Liverpool and transboundary partners to hold each other to account as they strengthen existing relationships, and form new ones.

## Data Availability

Anonymised data can be shared at request. Due to the nature of the research, the transcripts would have to be significantly redacted to remain anonymous as the data contains potentially identifying information.

## Acknowledgment

We would like to acknowledge all the partners that gave their time and thoughtful insight to shape this research and co-develop the principles presented. We thank Professor Frances Cowan and Dr. Webster Mavhu from LSTM and the Centre for Sexual and HIV and AIDS Research, Zimbabwe (CeSHHAR), Dr. Ndekya M. Oriyo, National Institute for Medical Research (NIMR) and Profesor Asma El Sony (Epi-Lab) for their critical comments and for the other valued, anonymous inputs we received. Thank you for administrative support for the ADAPT team at LSTM: Beth Hollihead, Faye Moody and Tracy Owen.

## Appendix 1: LSTM Partnership Survey – 2021

**Equitable partnerships: Amplifying perspectives from the countries where LSTM works**

**Introduction to the equitable partnership survey**

You have been invited to complete this survey because LSTM wants to understand how to better support equitable partnerships in the countries where it works. If you choose to participate, your views will be instrumental in shaping and informing LSTM’s practice and strategic vision in this area to ensure all our partnerships are of mutual benefit.

We have identified a number of topics for investigation (these are are adapted from the Research Fairness initiative domains of: fairness of opportunity; fair process; fair sharing of benefits, costs and outcomes) but please add any other issues you would like to tell us about, using the free-text boxes. You also have an opportunity to include your contact details at the end if you wish to be contacted for a follow up interview – however, if you wish to remain anonymous in the survey, but have a particular interest in being involved in an individual interview please.

All data will be kept securely and limited members from [LSTM](https://www.lstmed.ac.uk/) & [LVCT Health](https://lvcthealth.org/) will have direct access to this. All your views expressed in this survey will be anonymous: we will anonymise data during analysis. We may analyse data by characteristics (e.g. gender, location of institution, age or type of activity of participant) but will ensure that in reporting no information that would reveal the identity of a respondent is used. By taking part you consent to us analysing this data.

We will report the findings of the survey to all LSTM partners through a report and also discuss the findings and potential ways to improve our partnership to be mutually beneficial. The survey should take no more than 30 minutes to complete.

Section 1: Characteristics

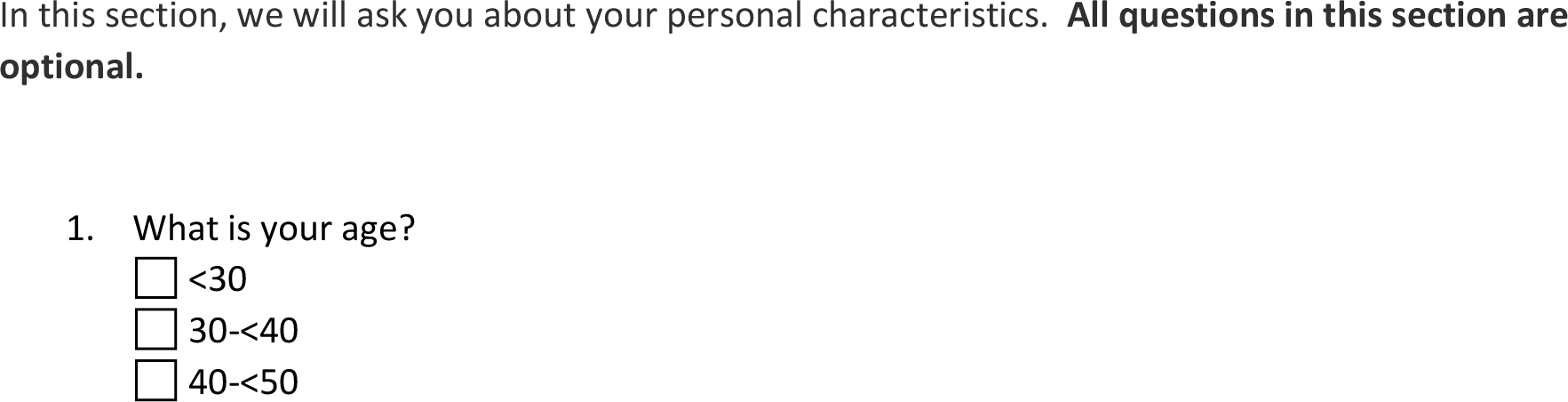
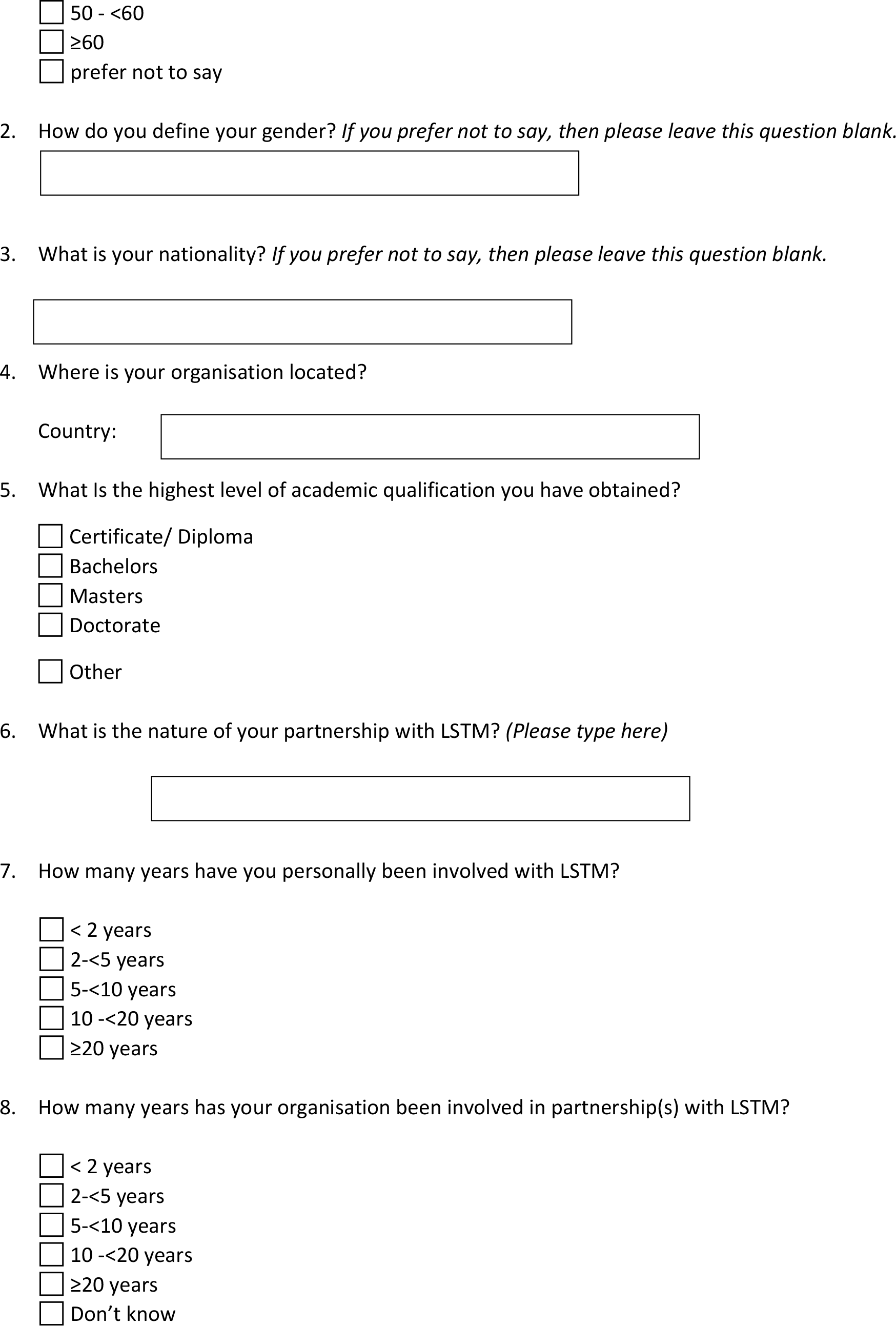
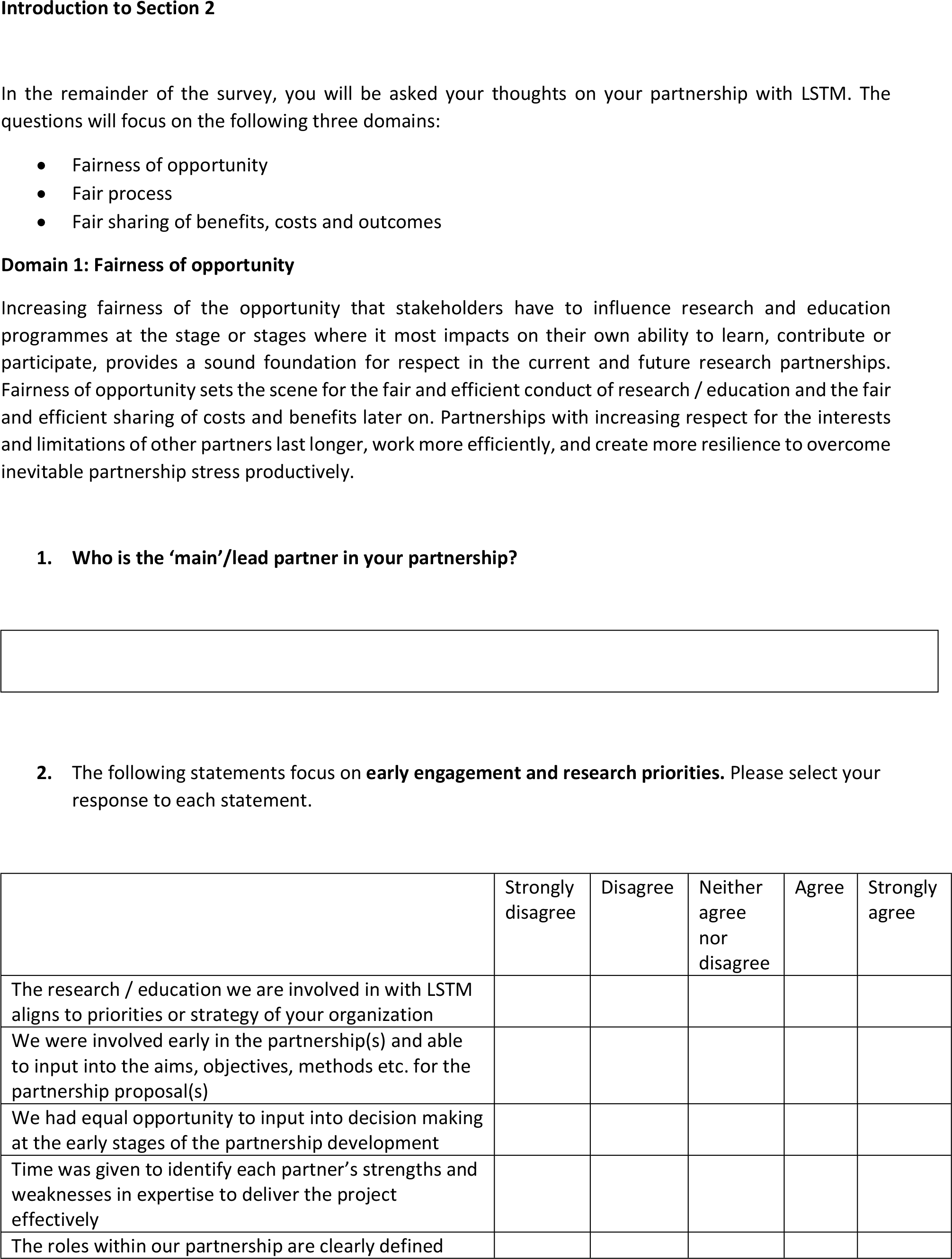
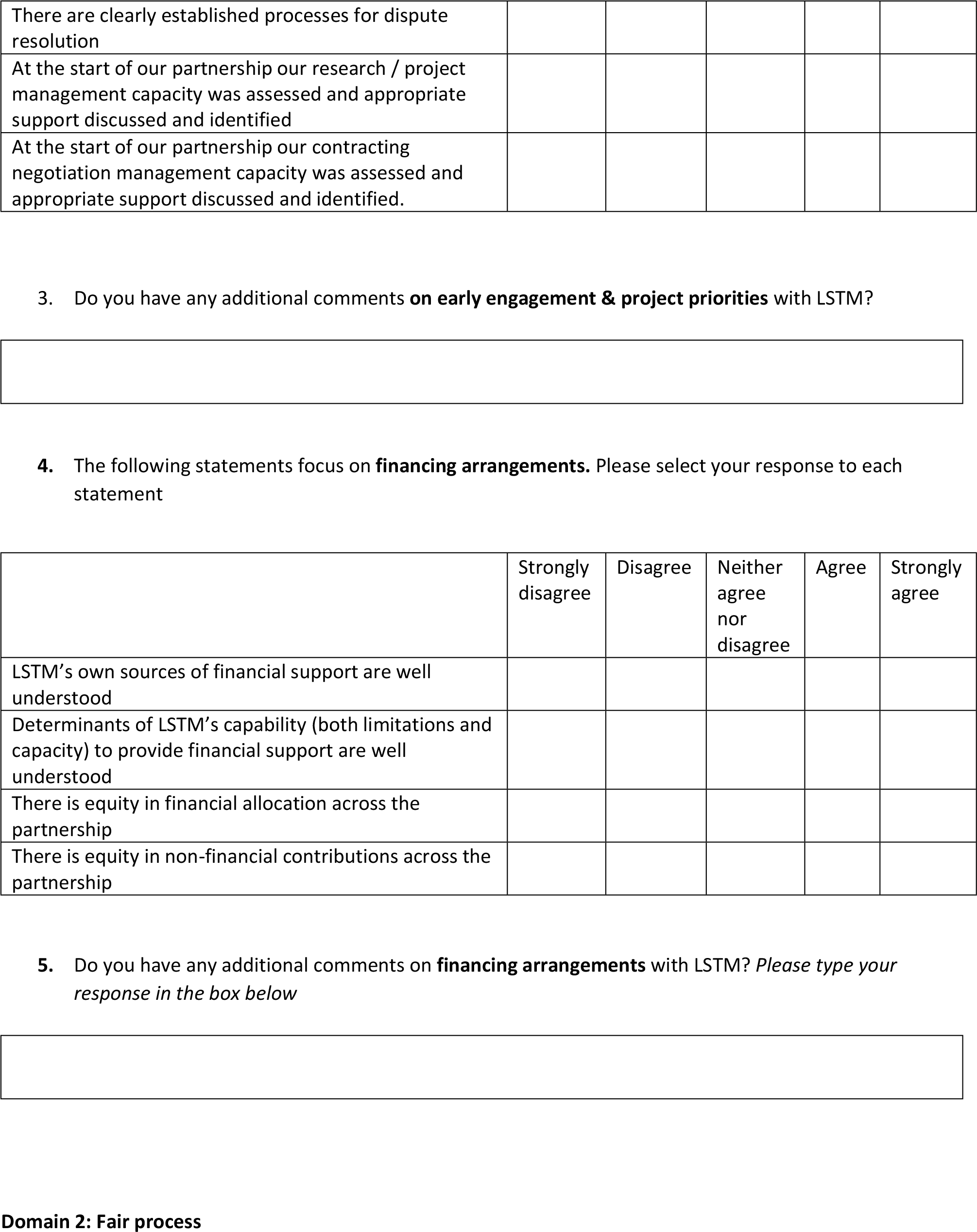
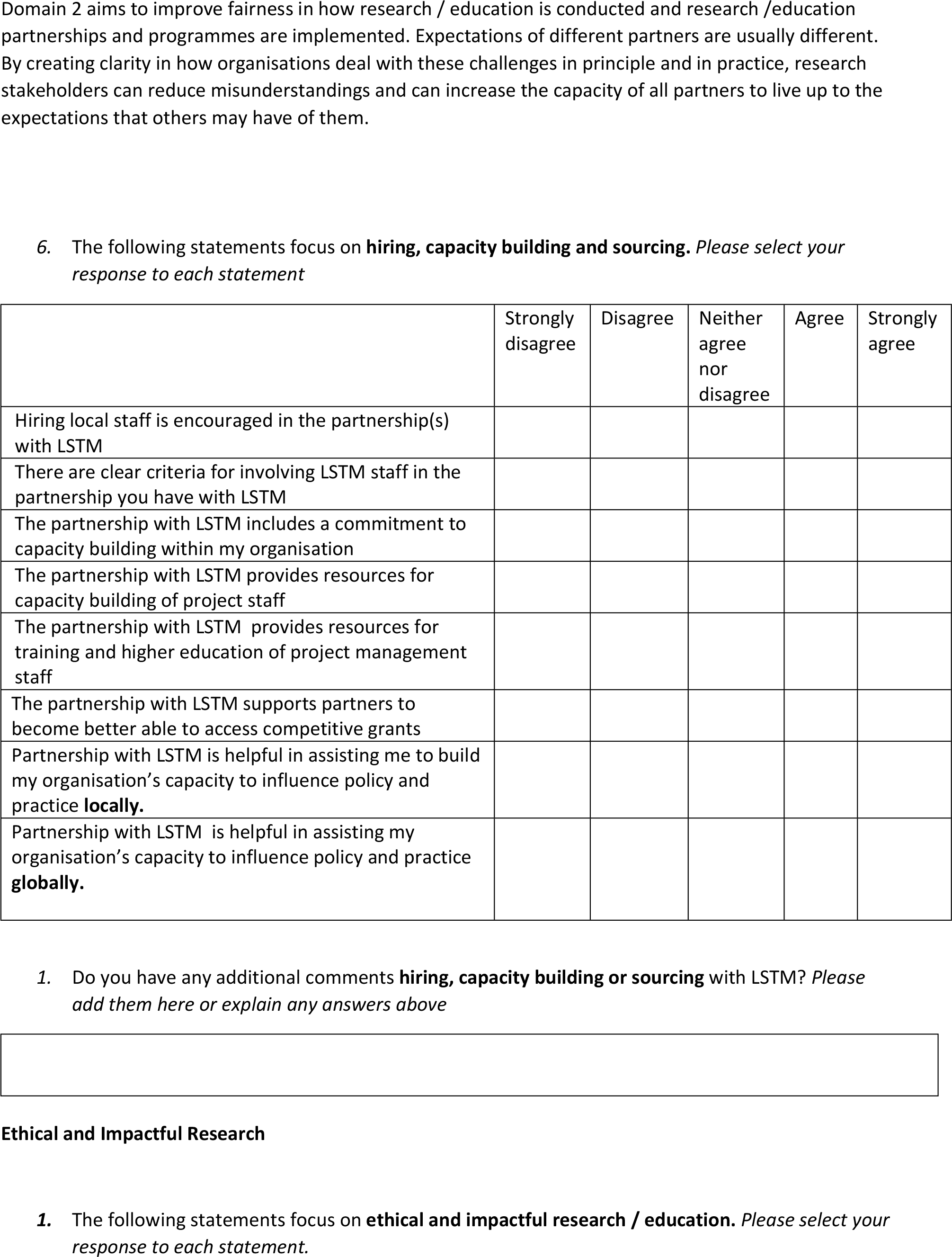
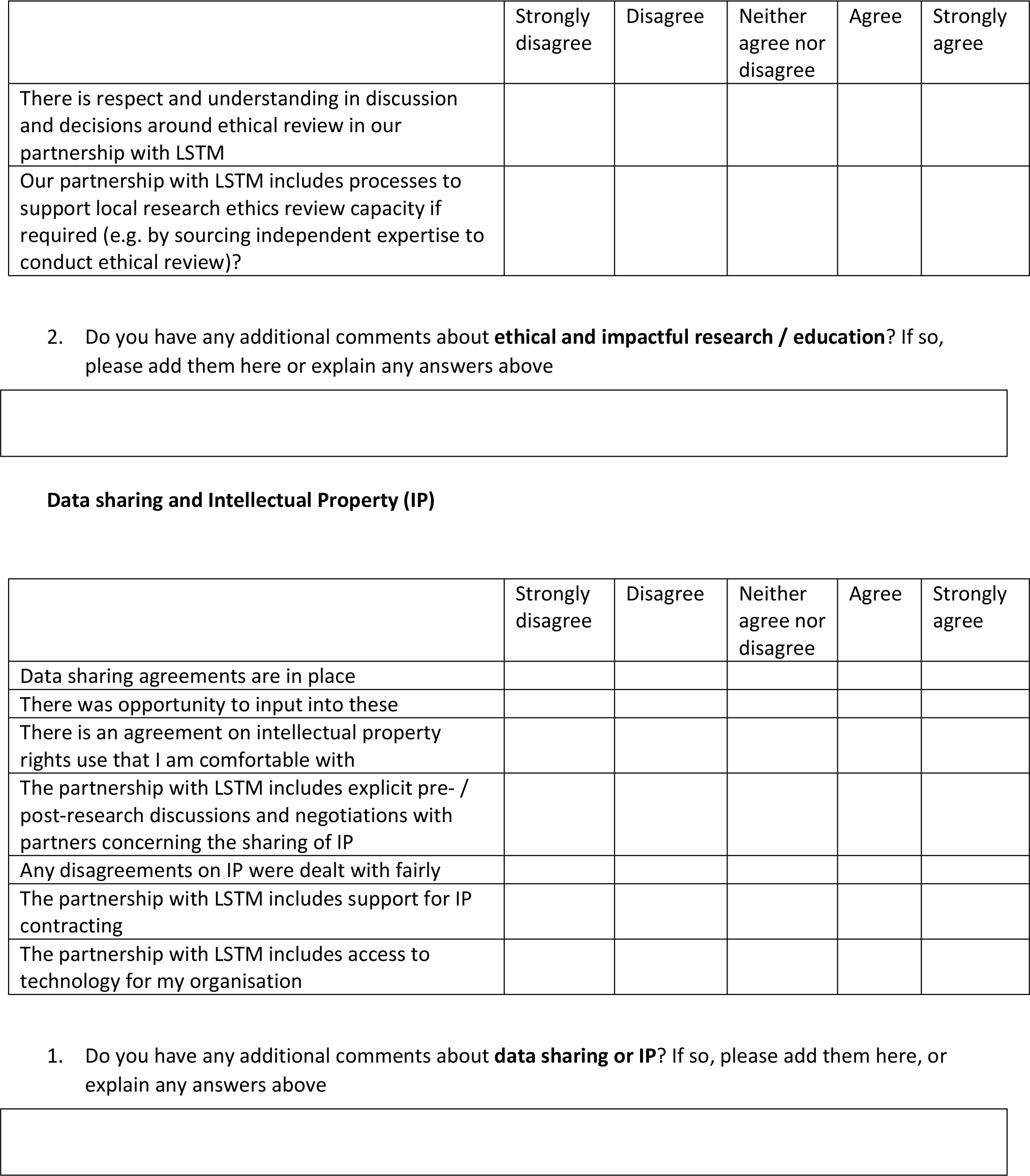
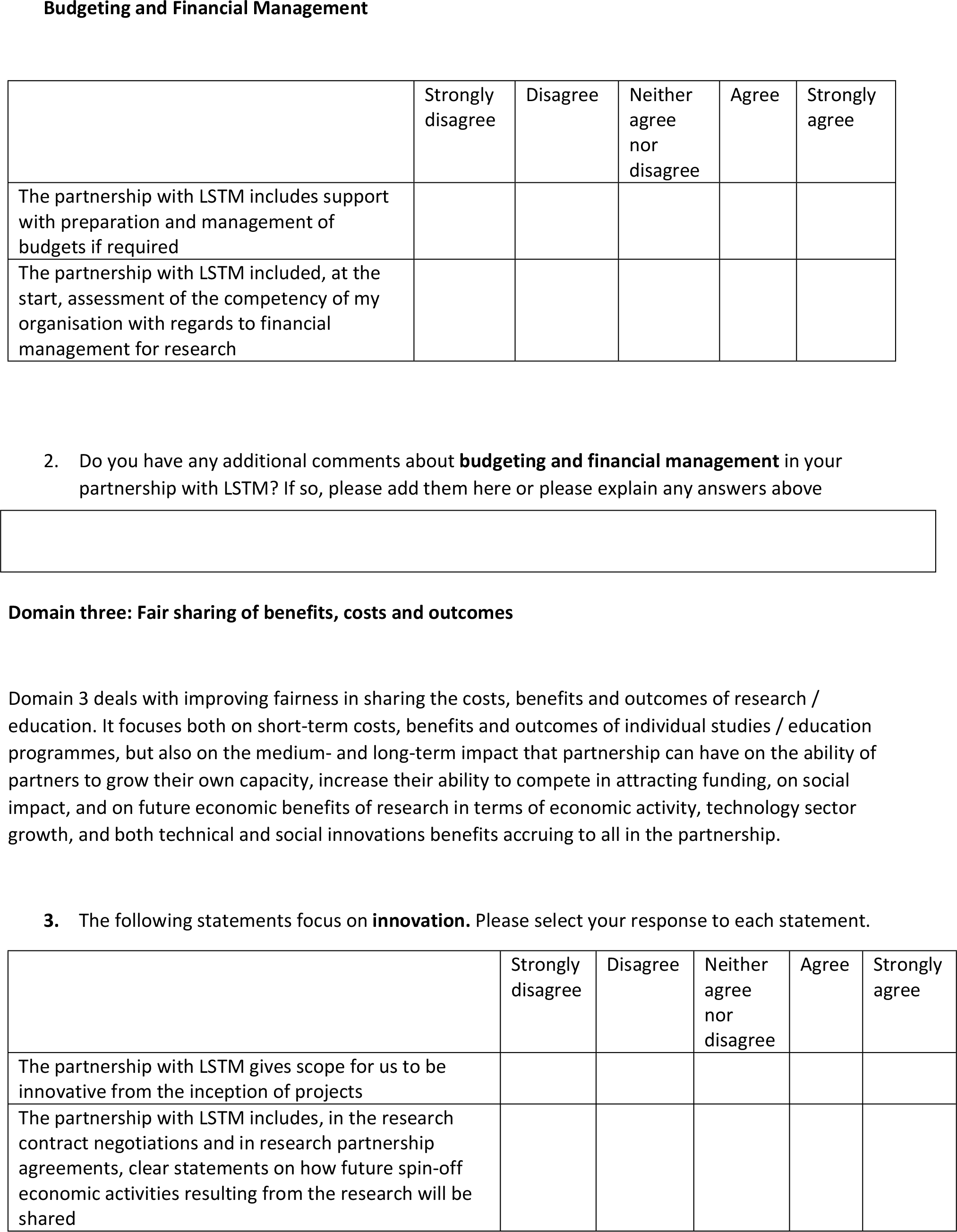
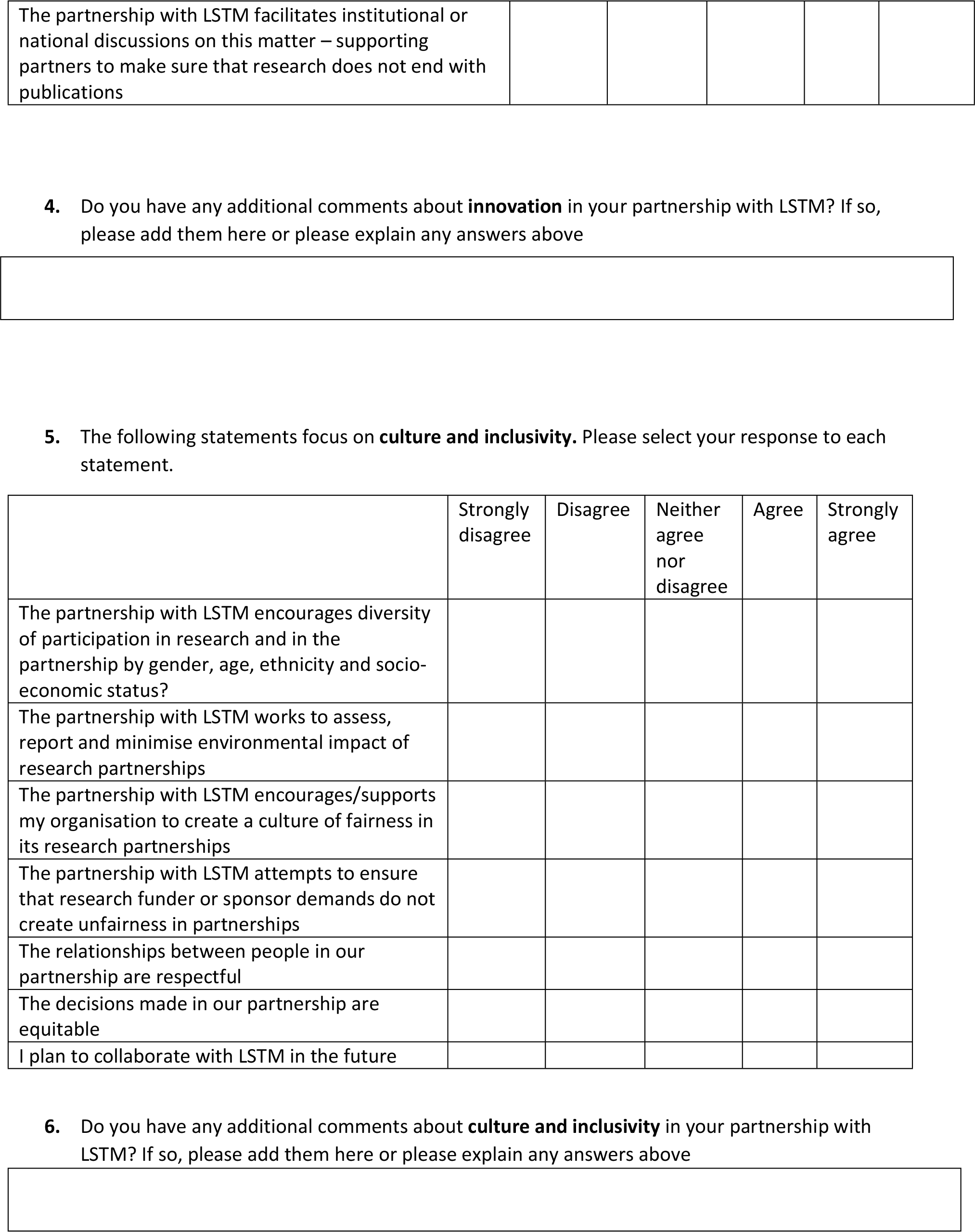
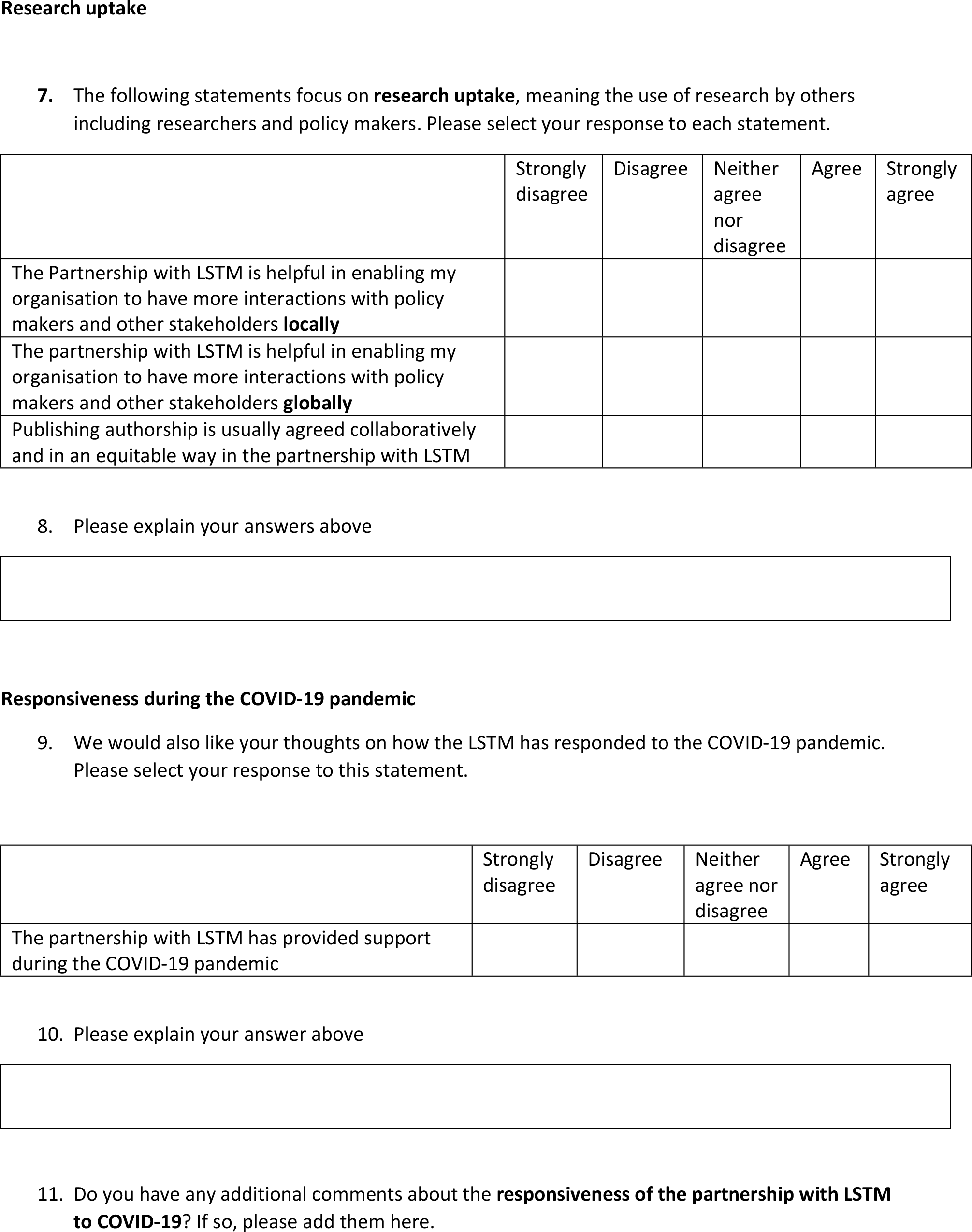
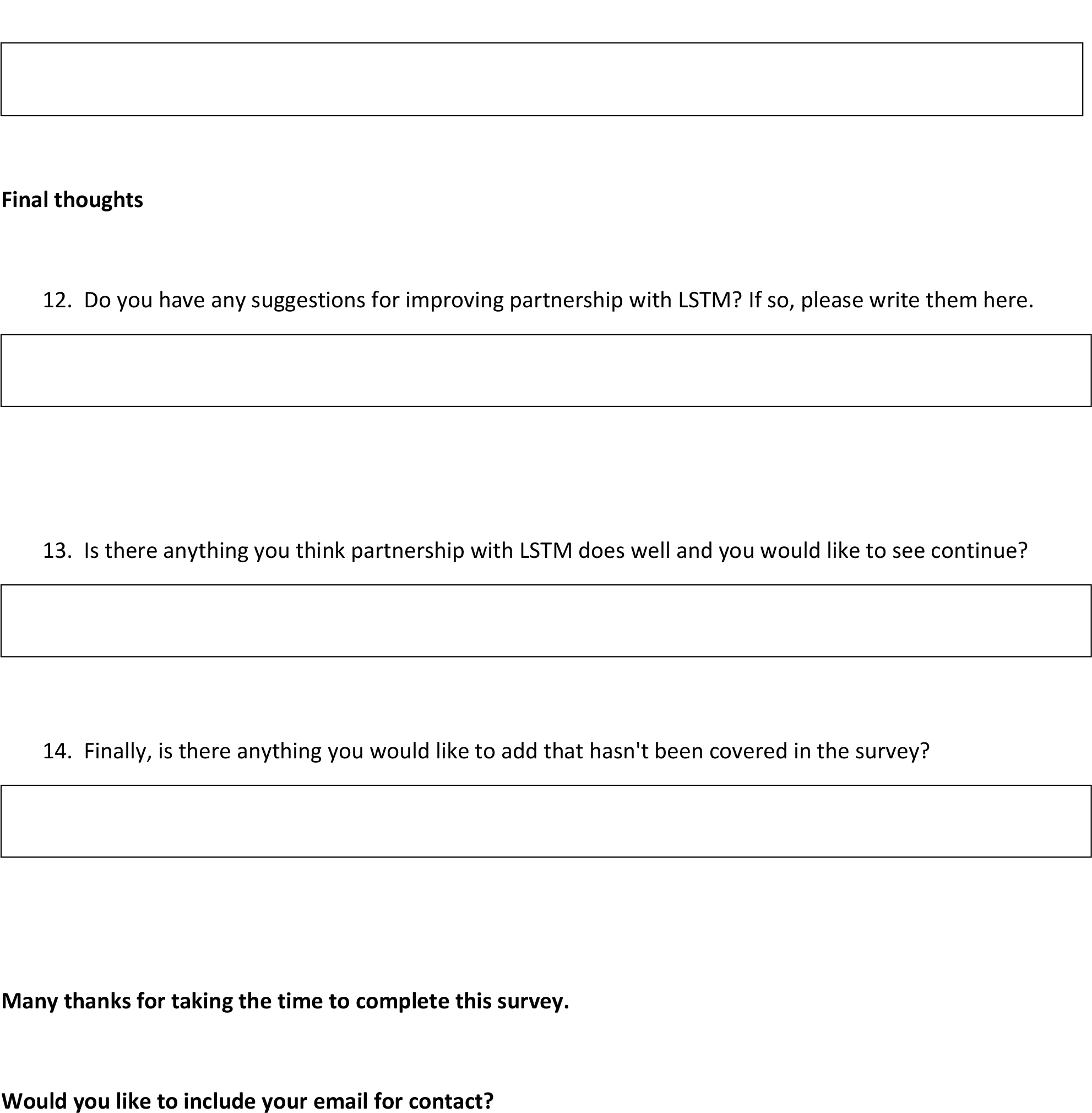

## Appendix 2: Abridged anonymised survey responses (does not include free text responses)

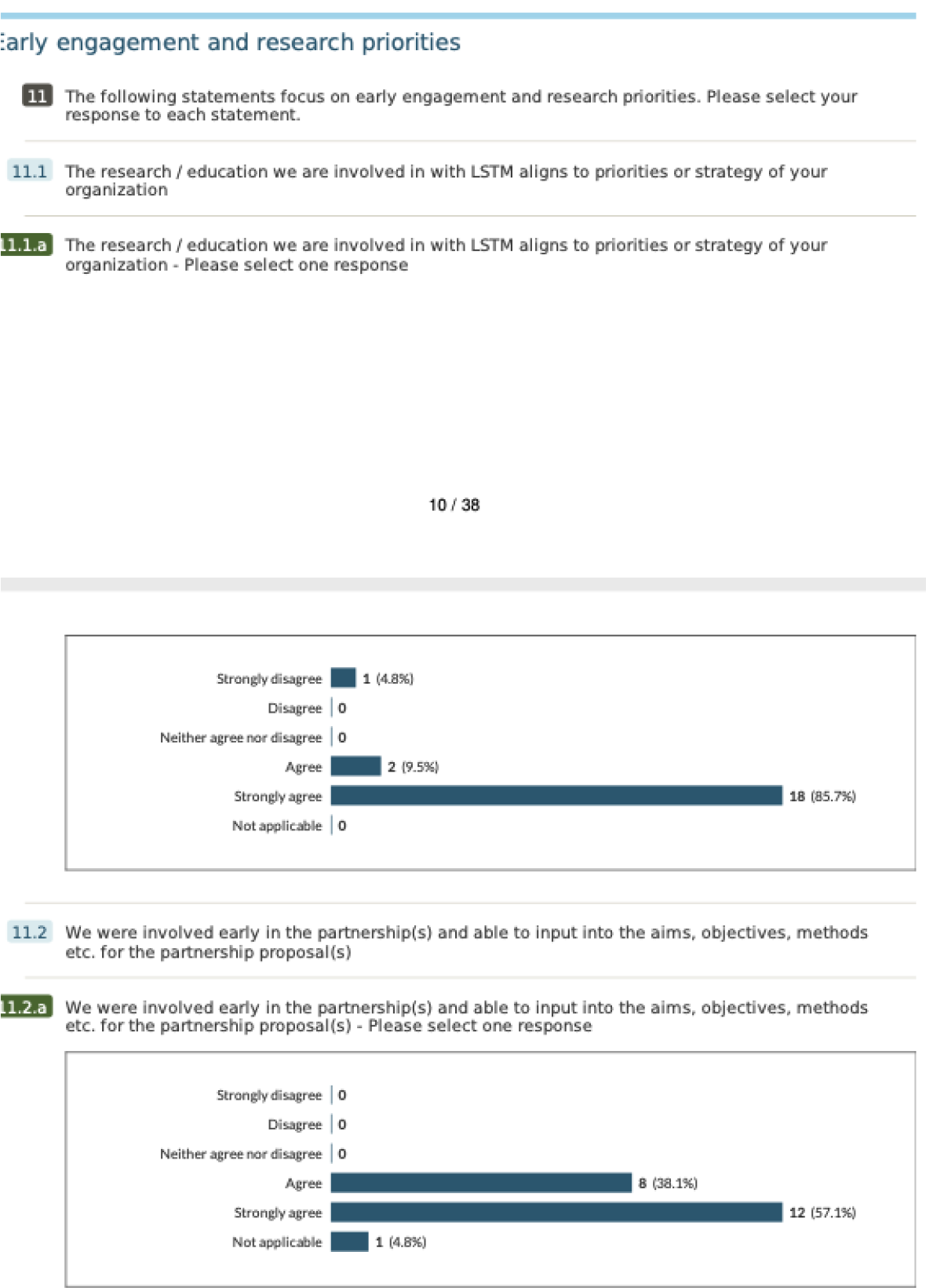
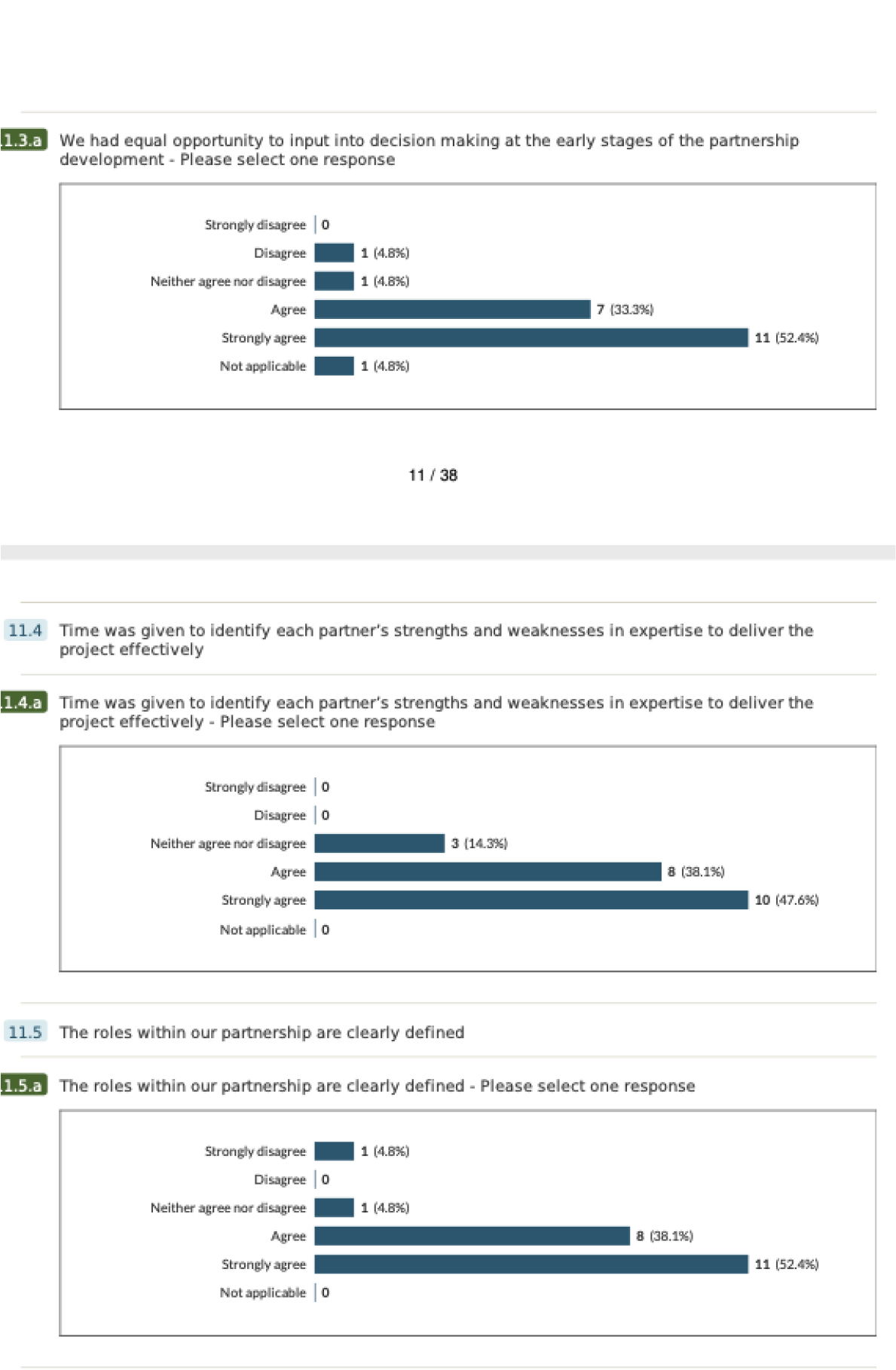
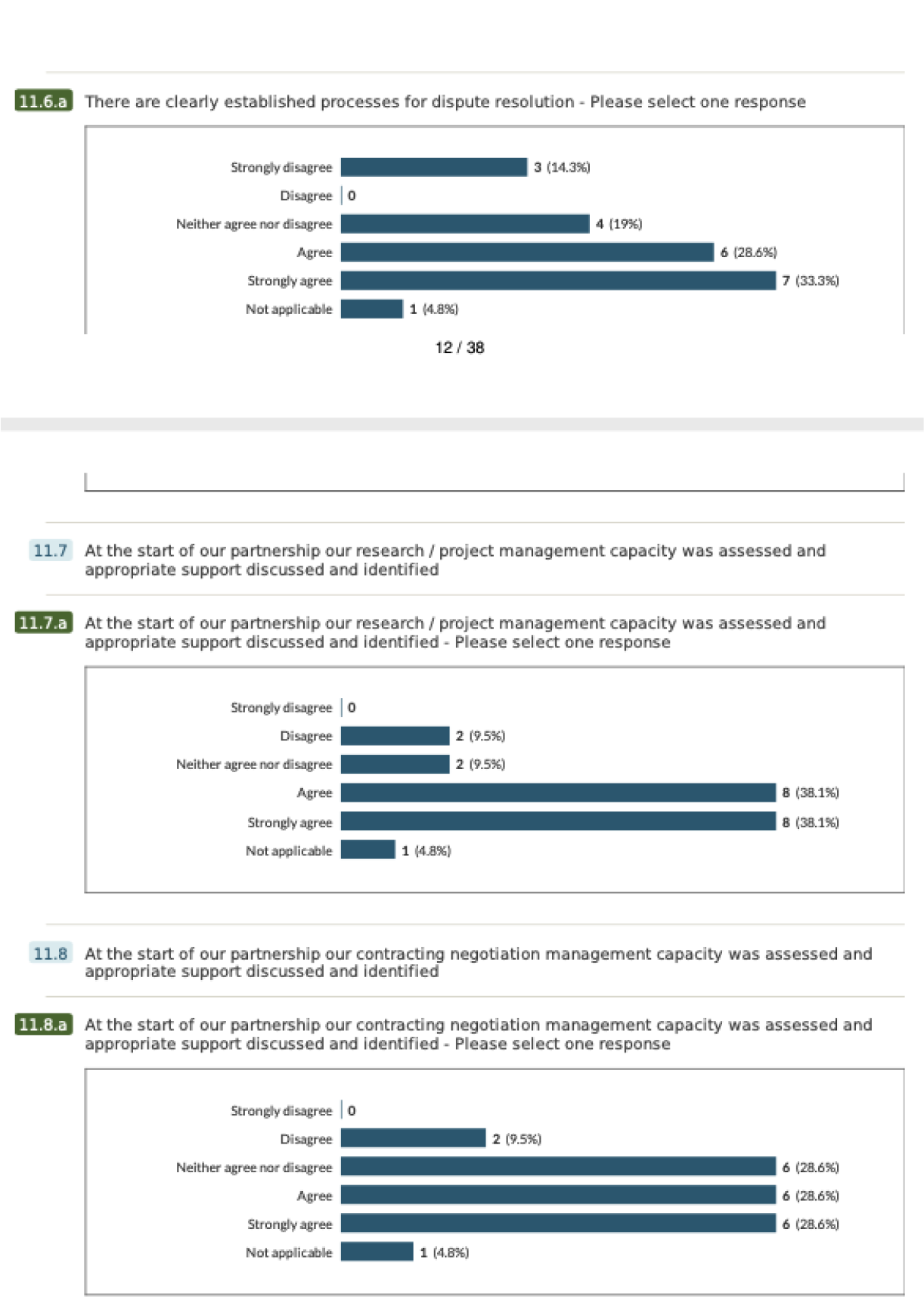
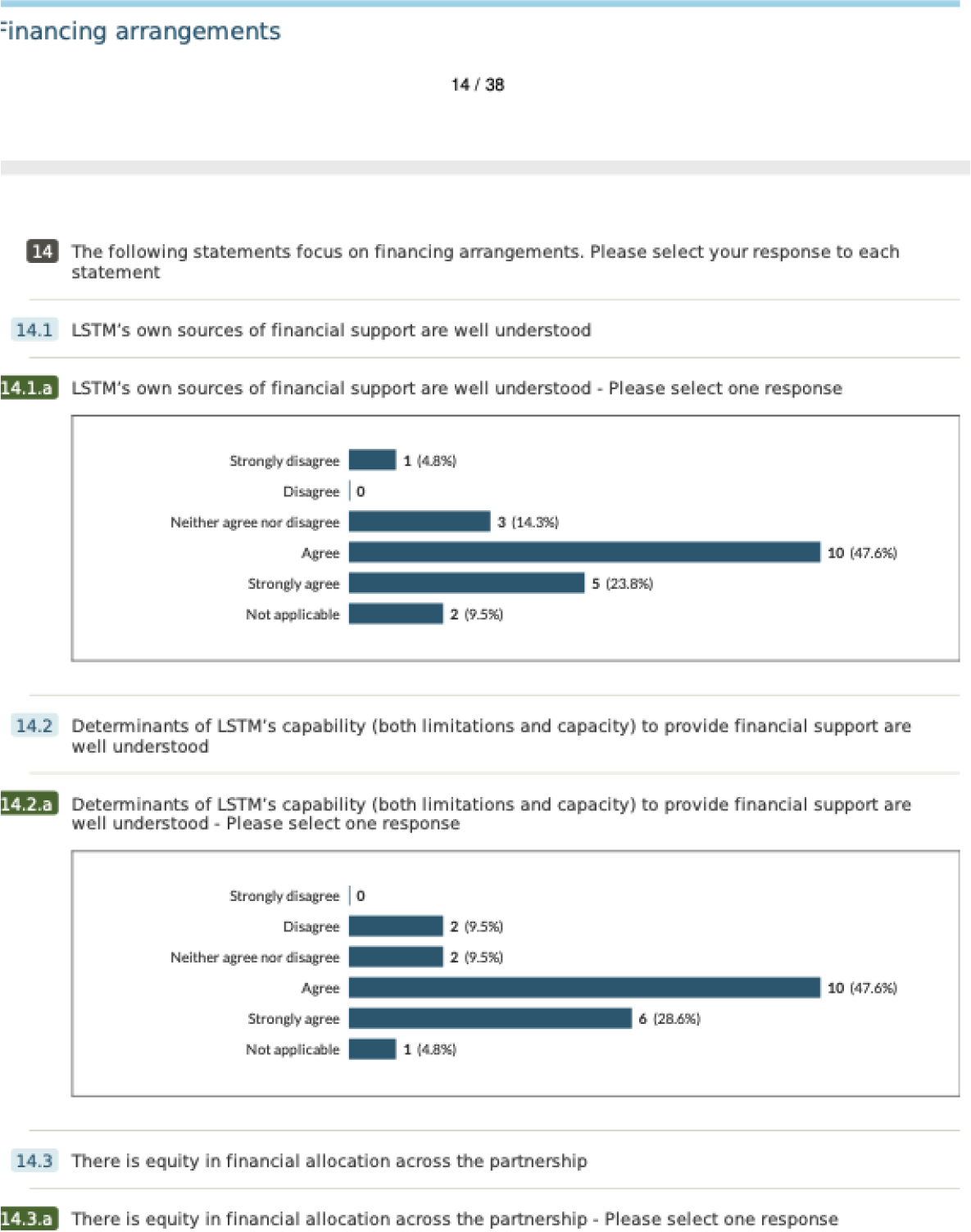
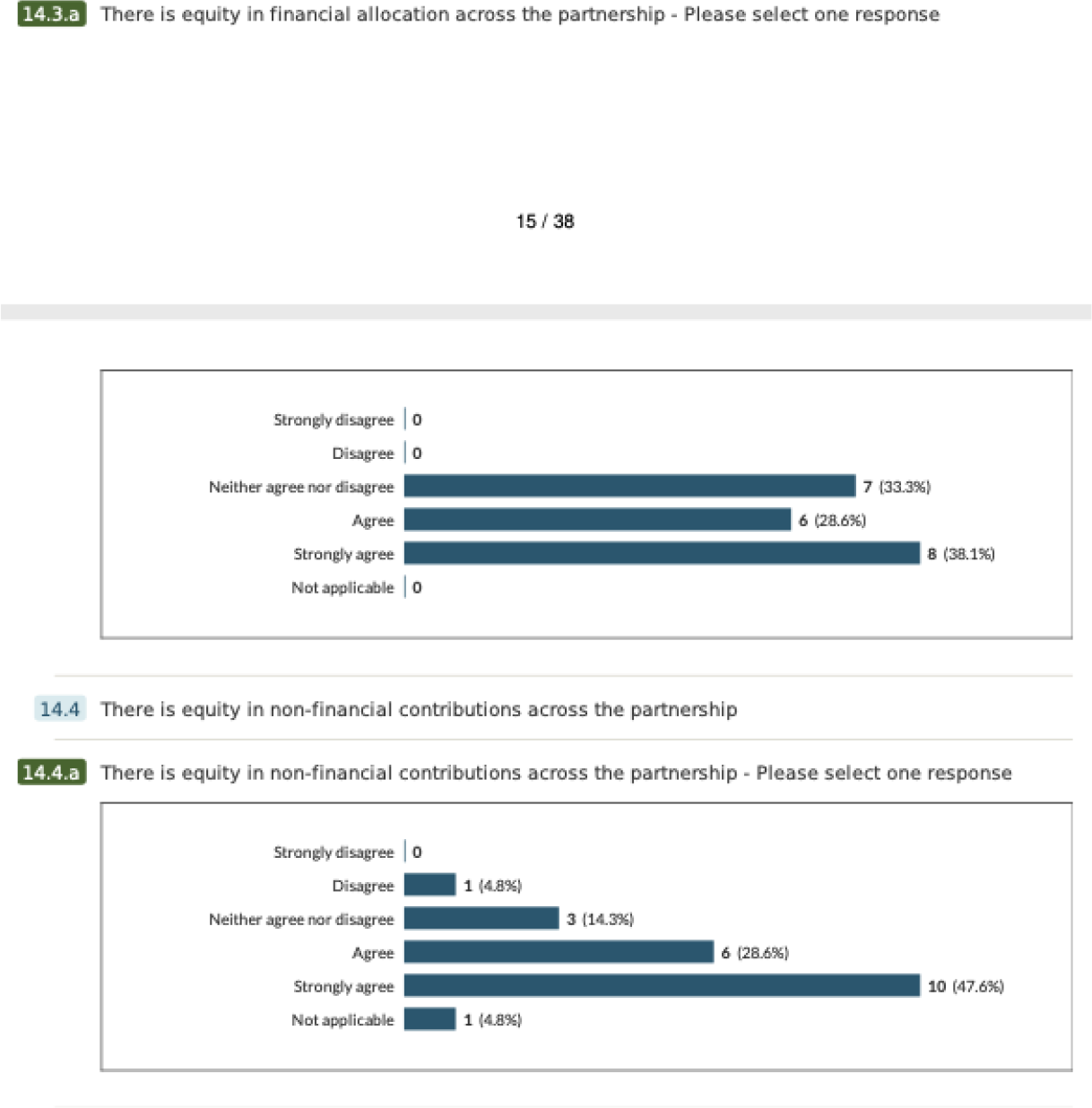
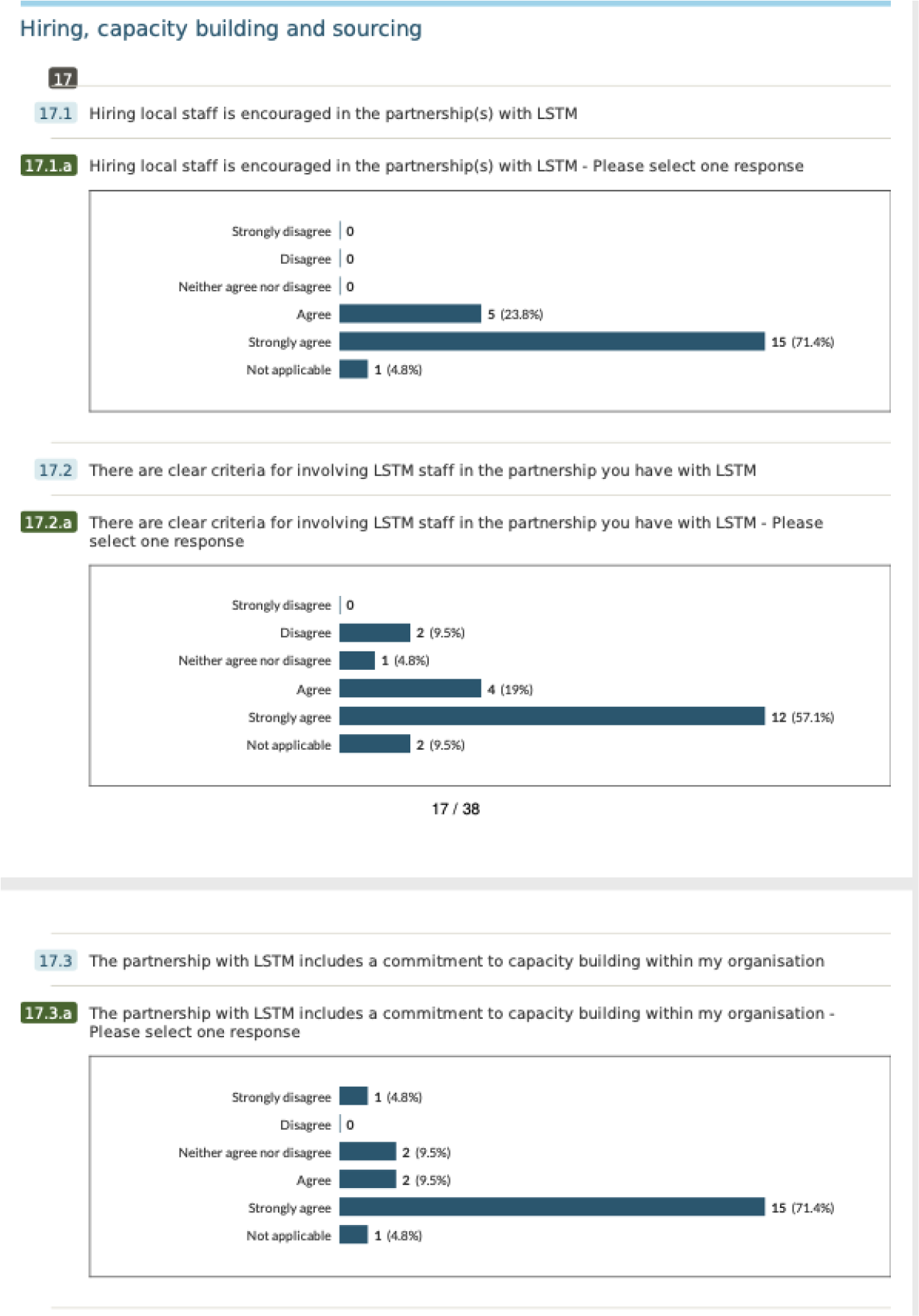
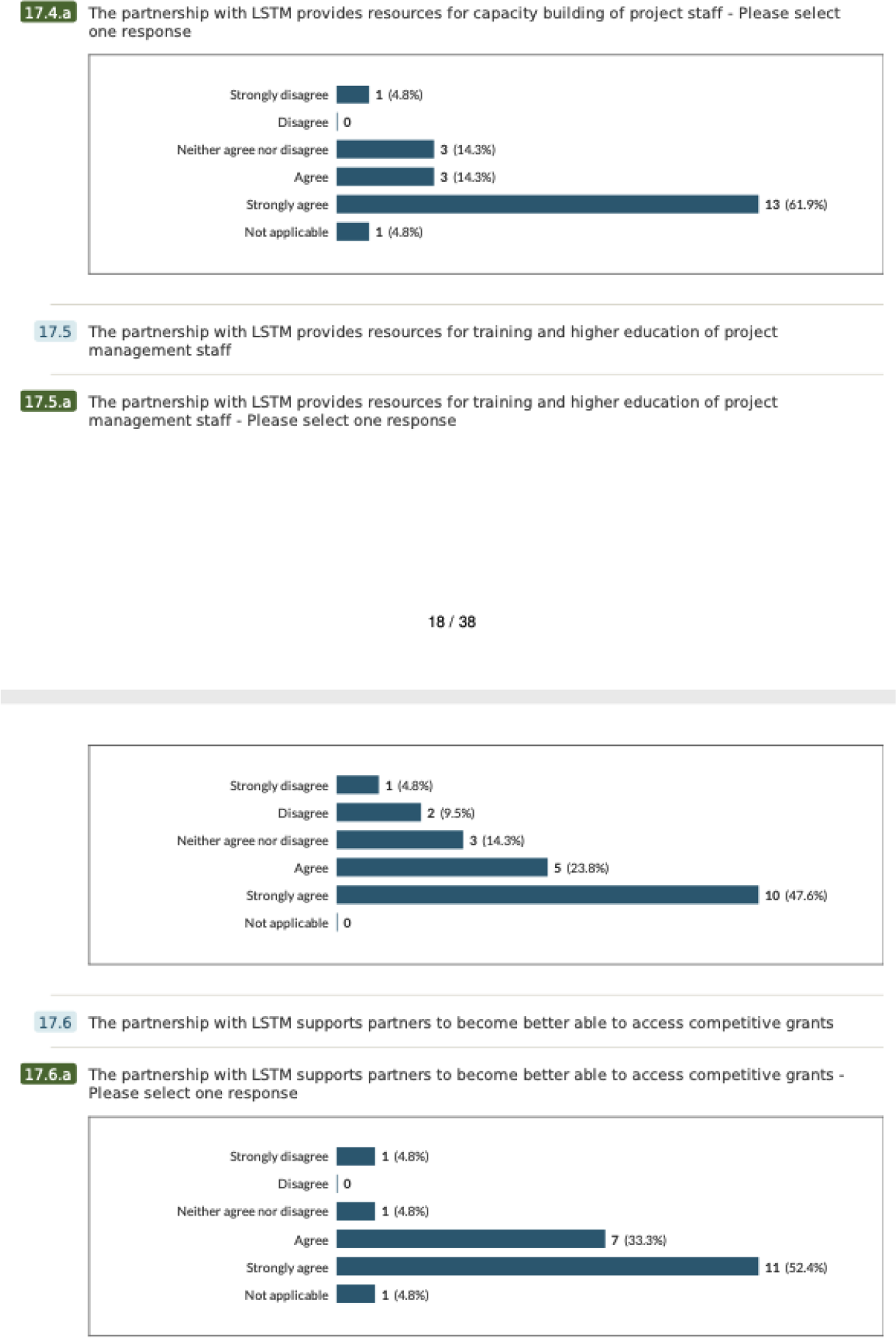
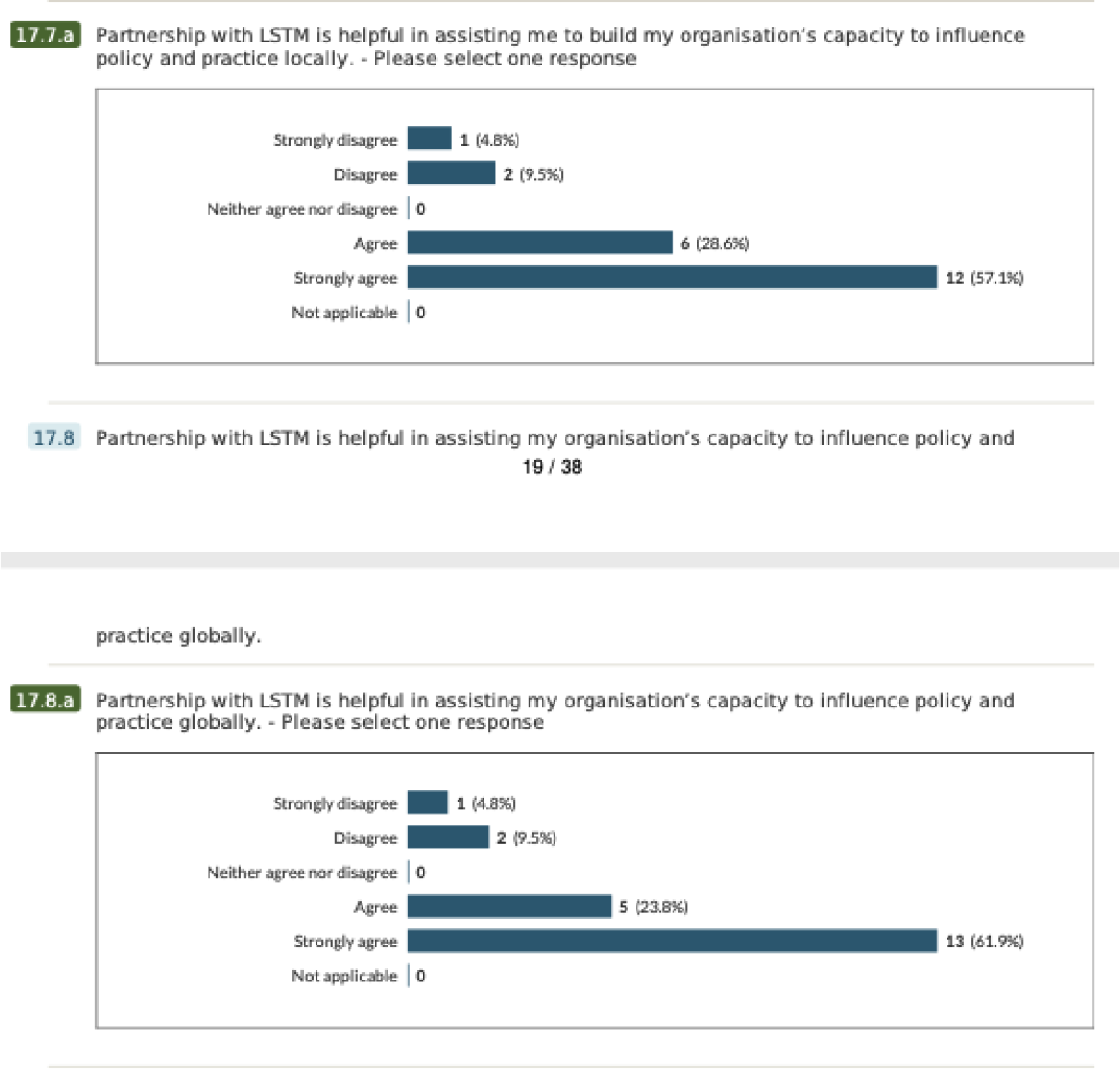
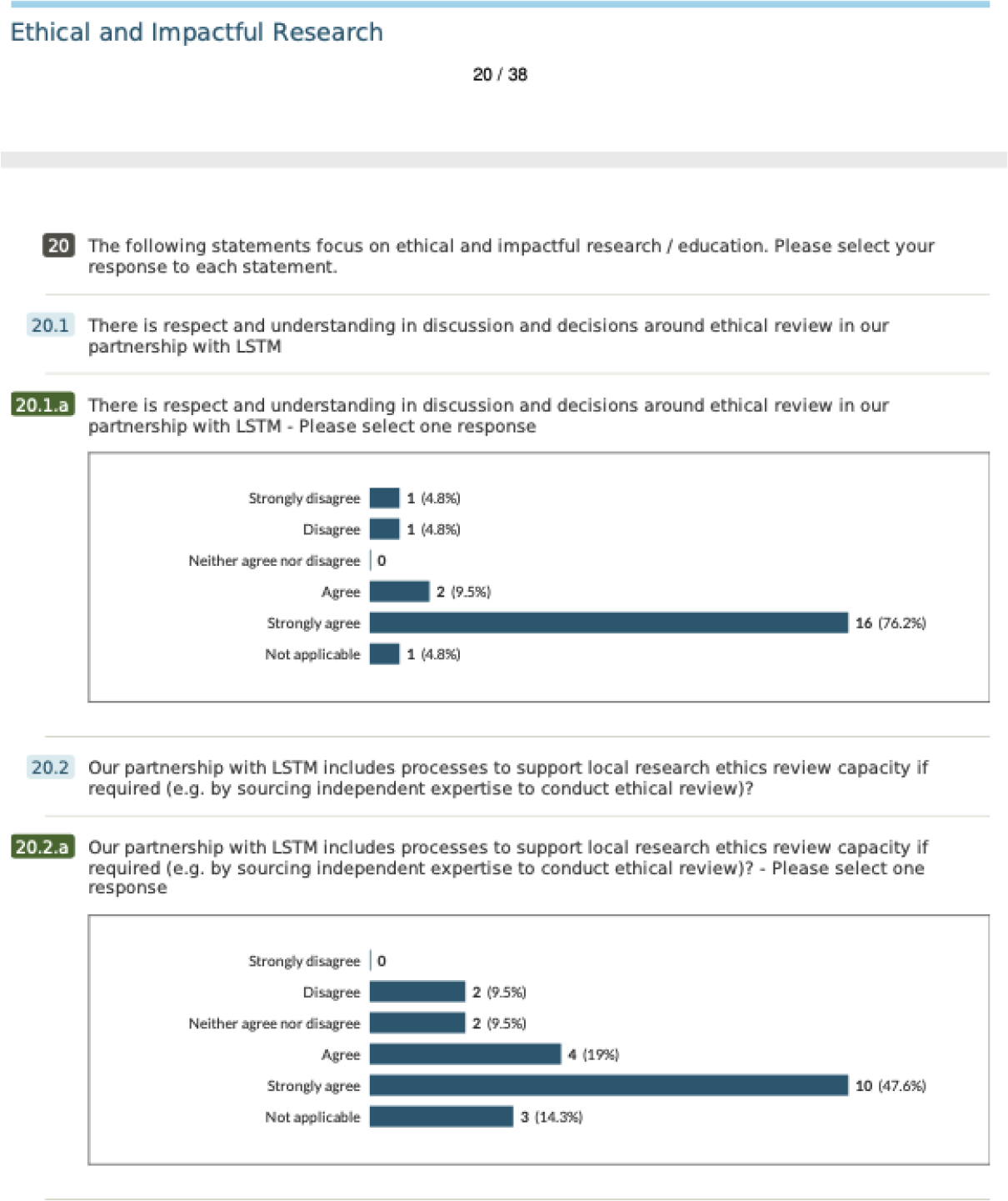
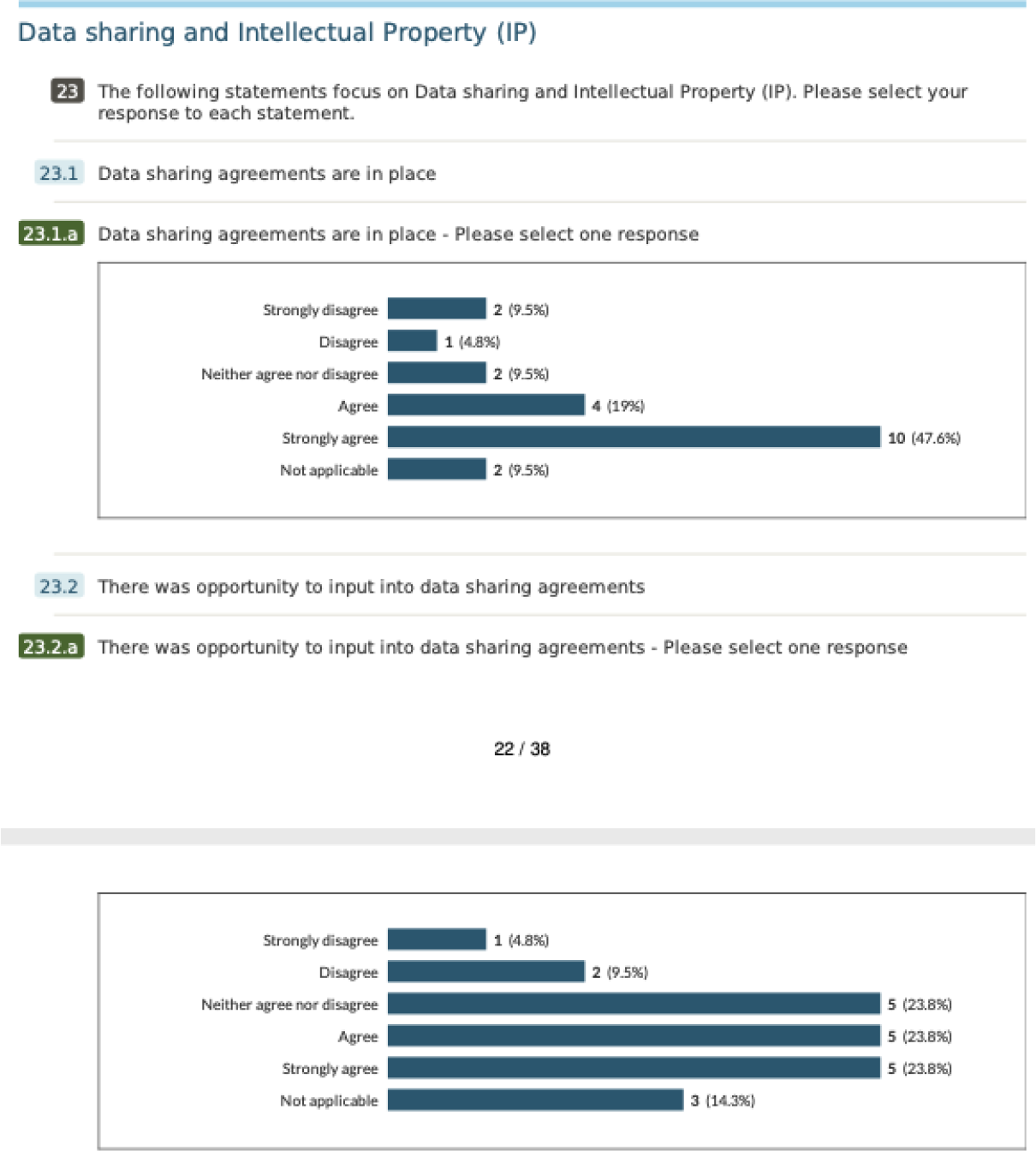
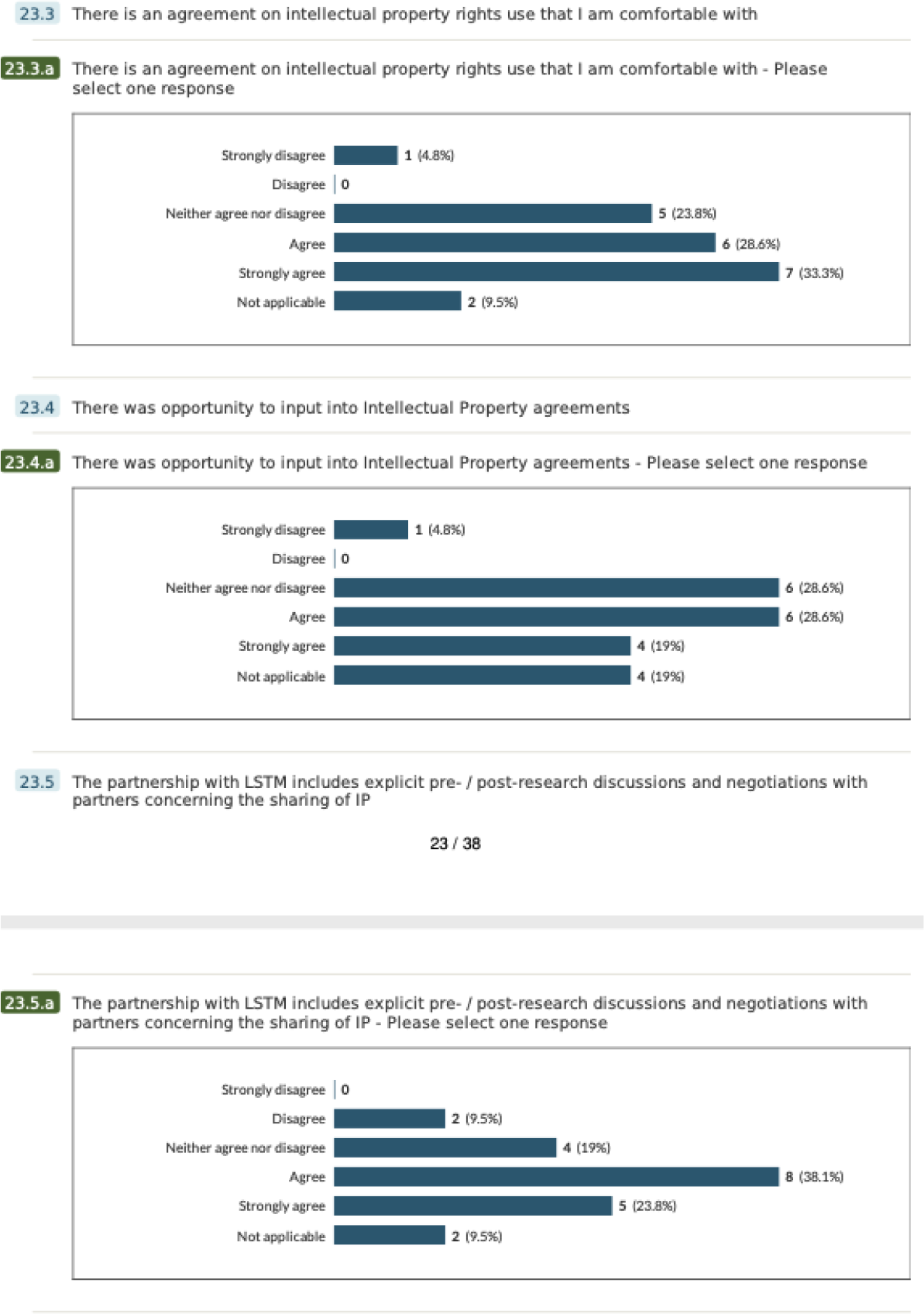
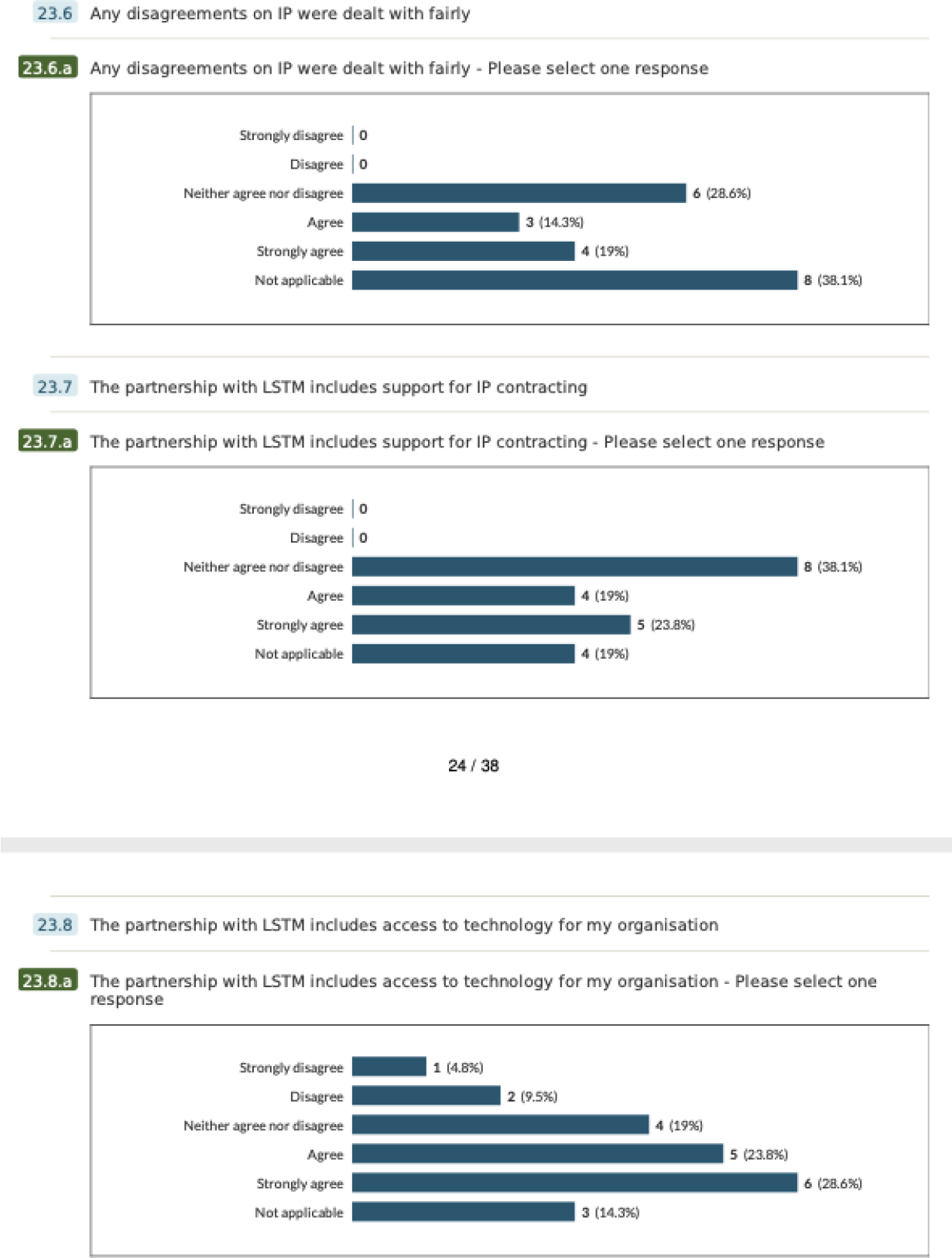
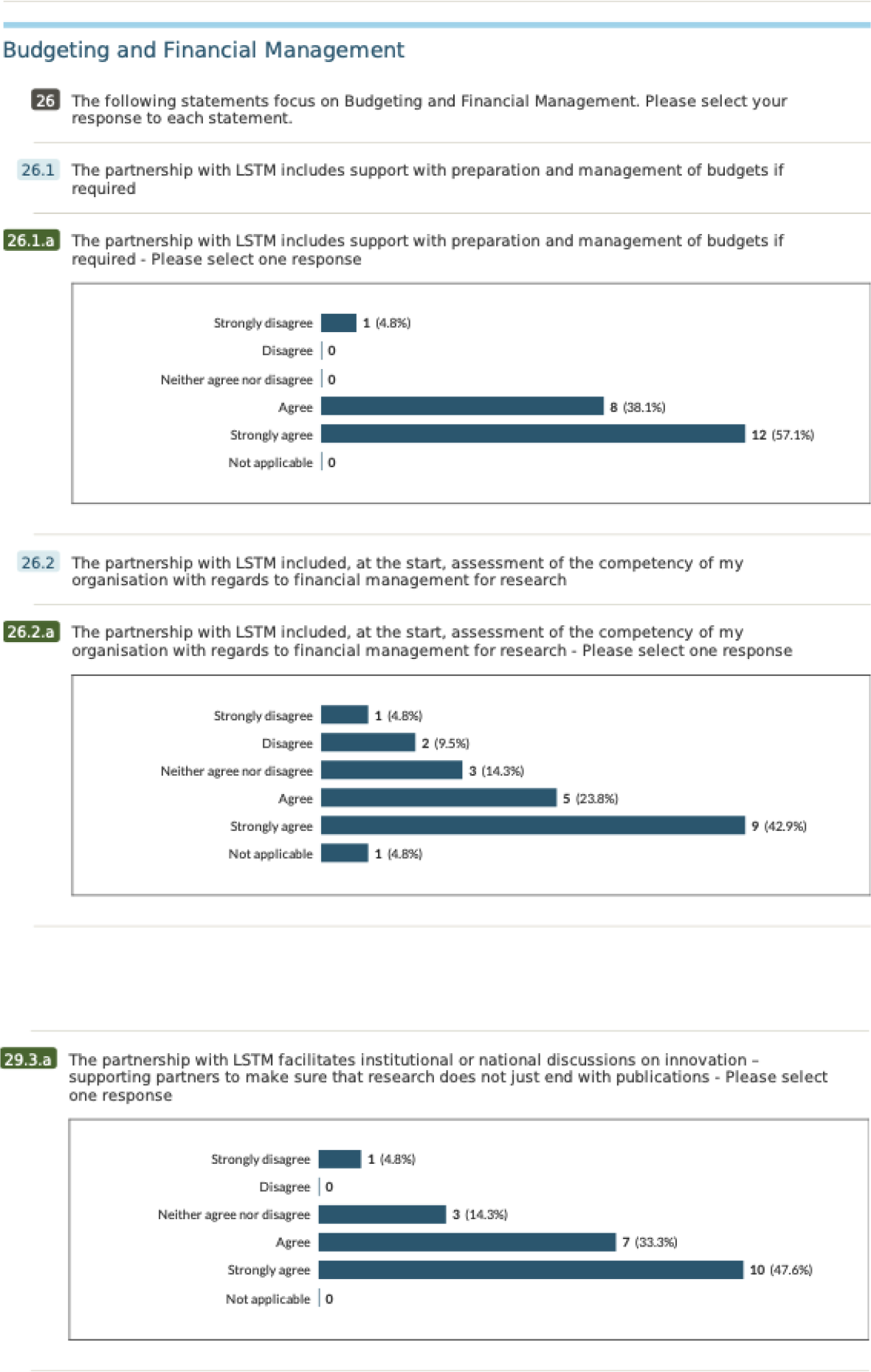
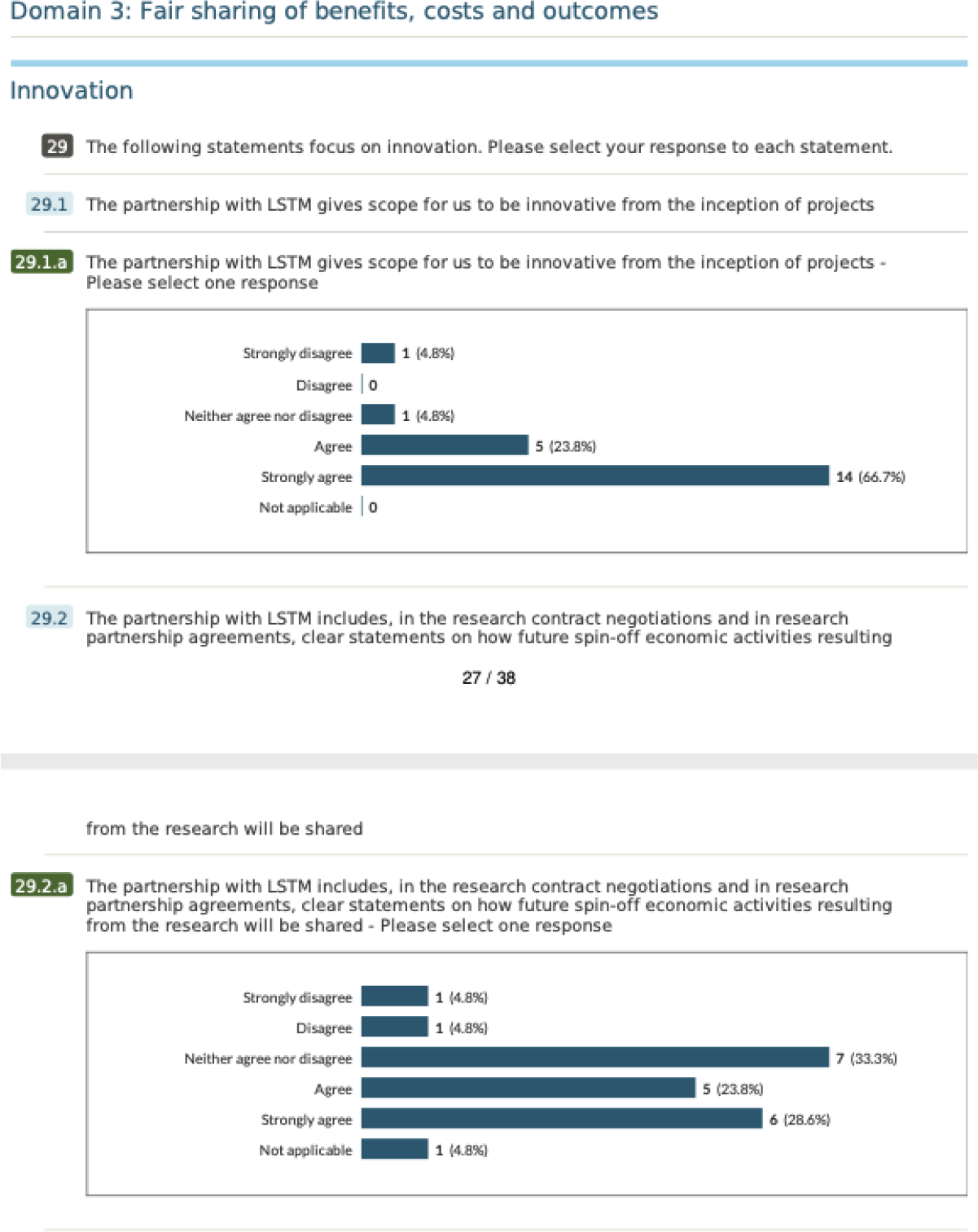
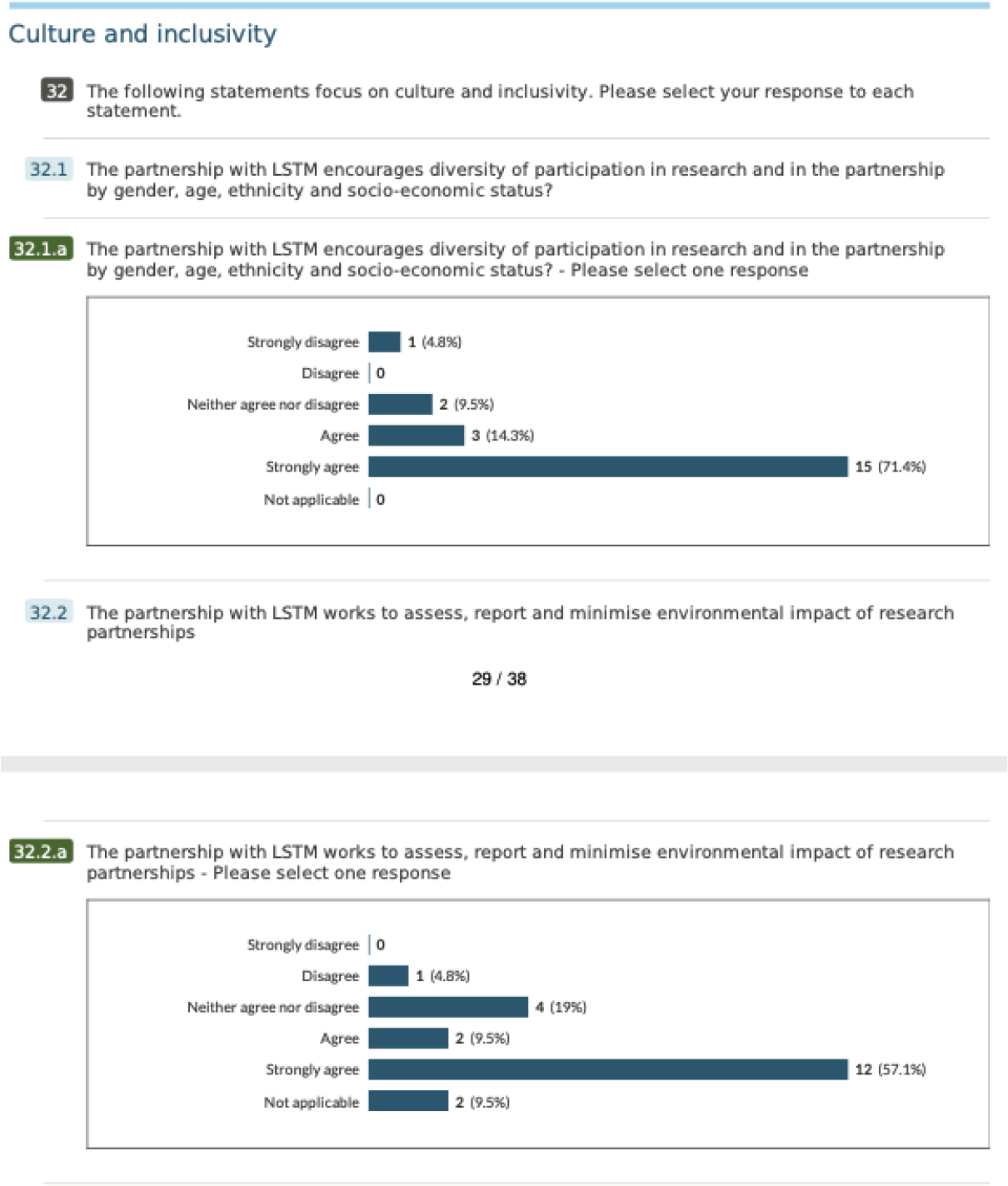
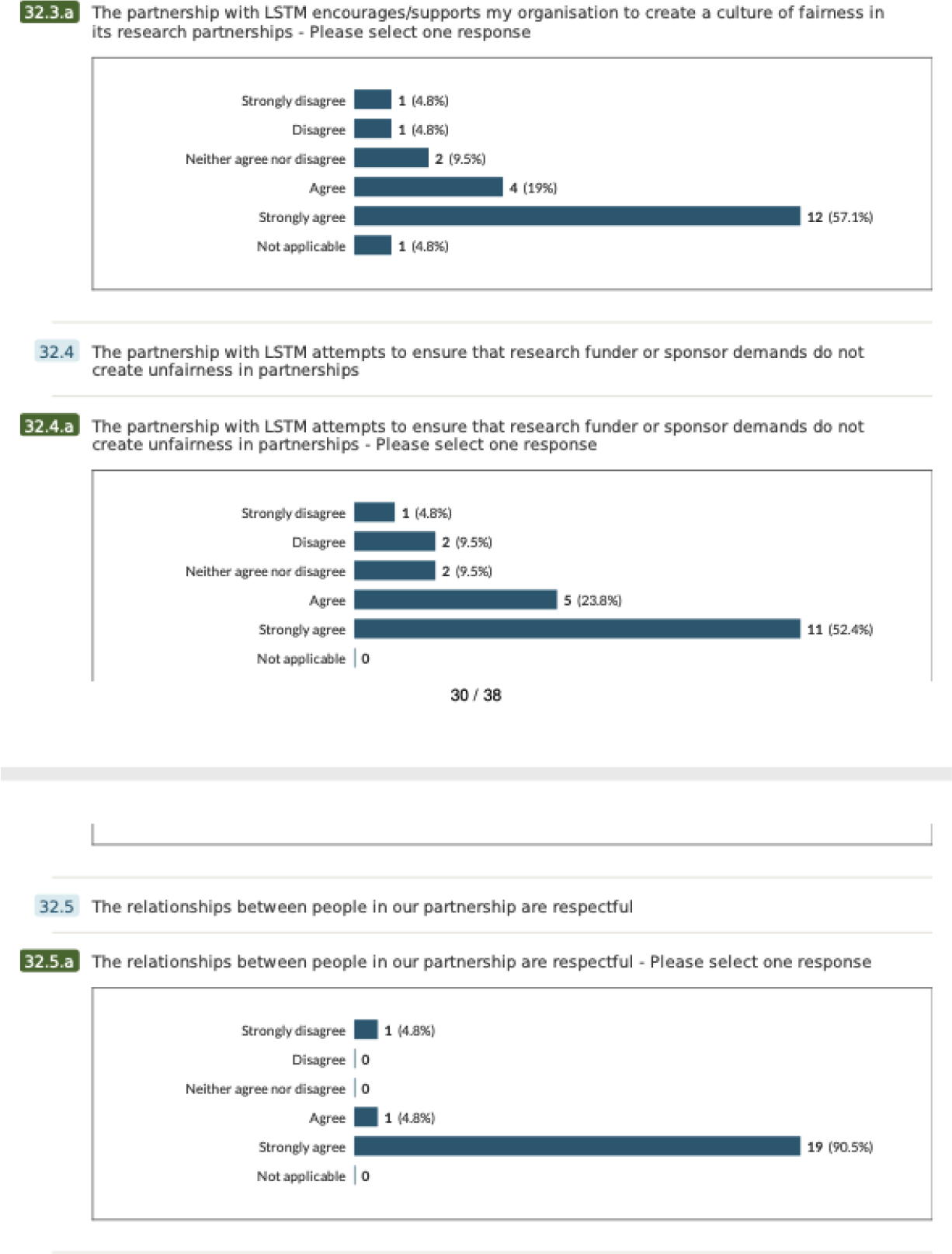
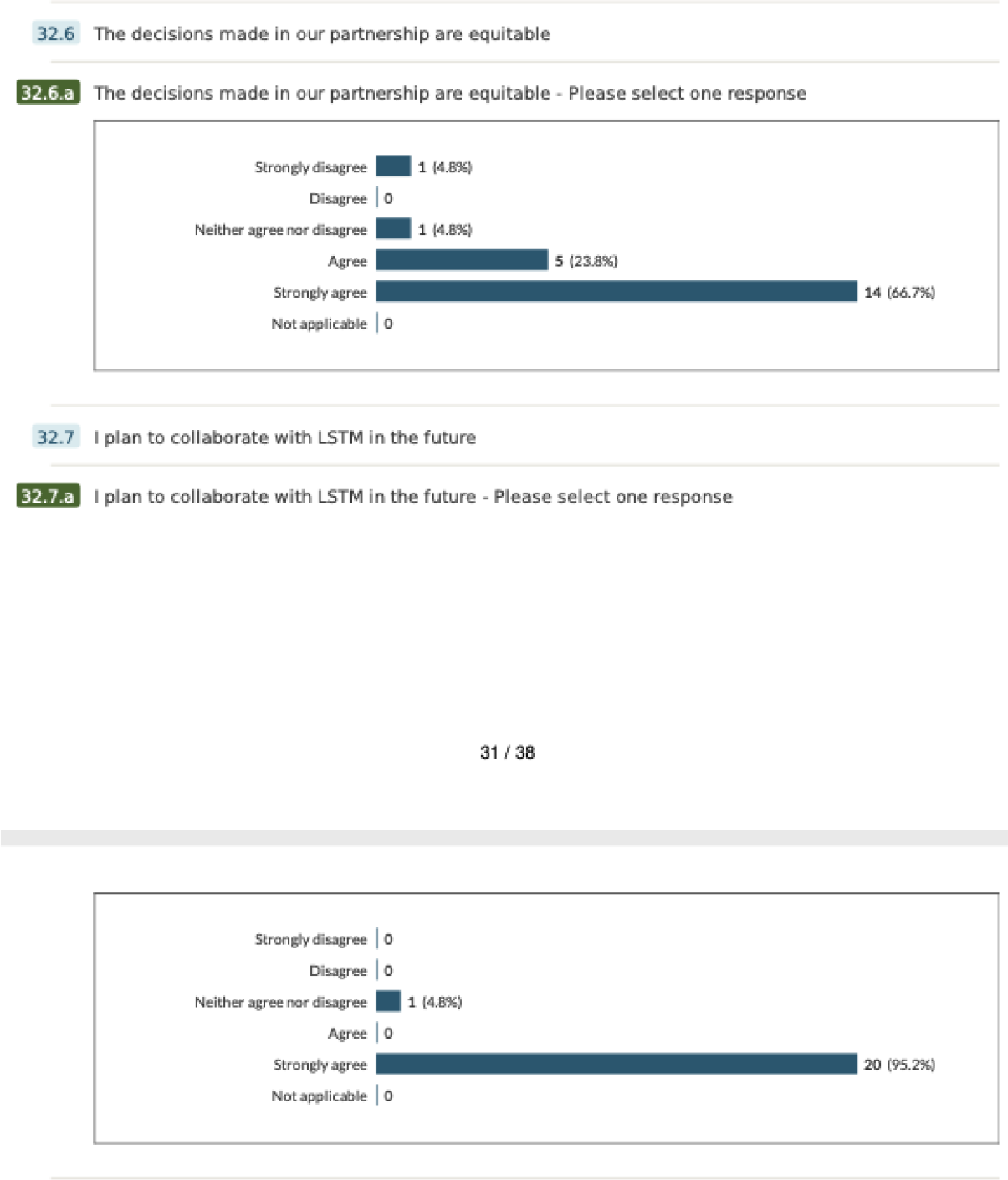
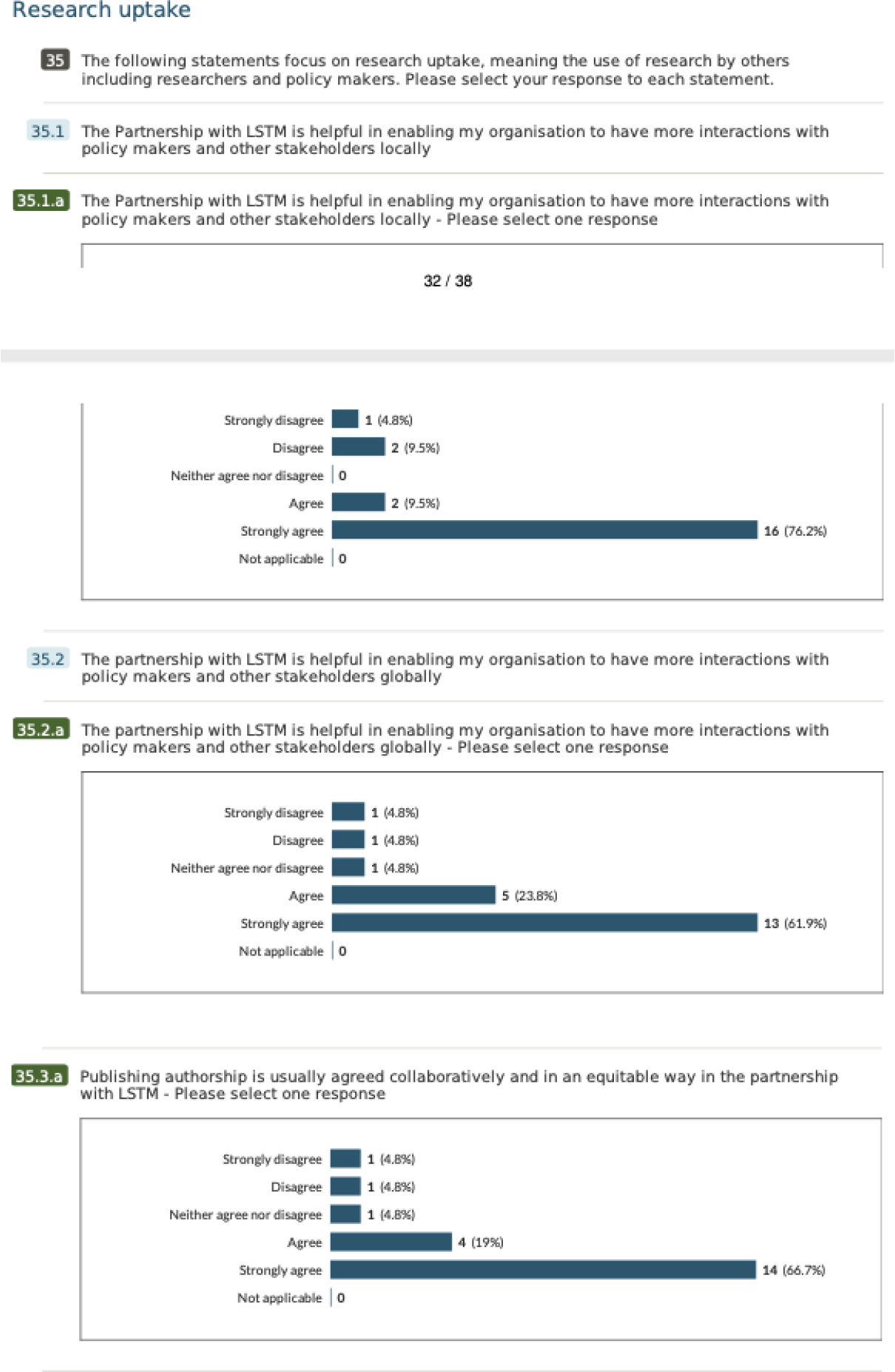
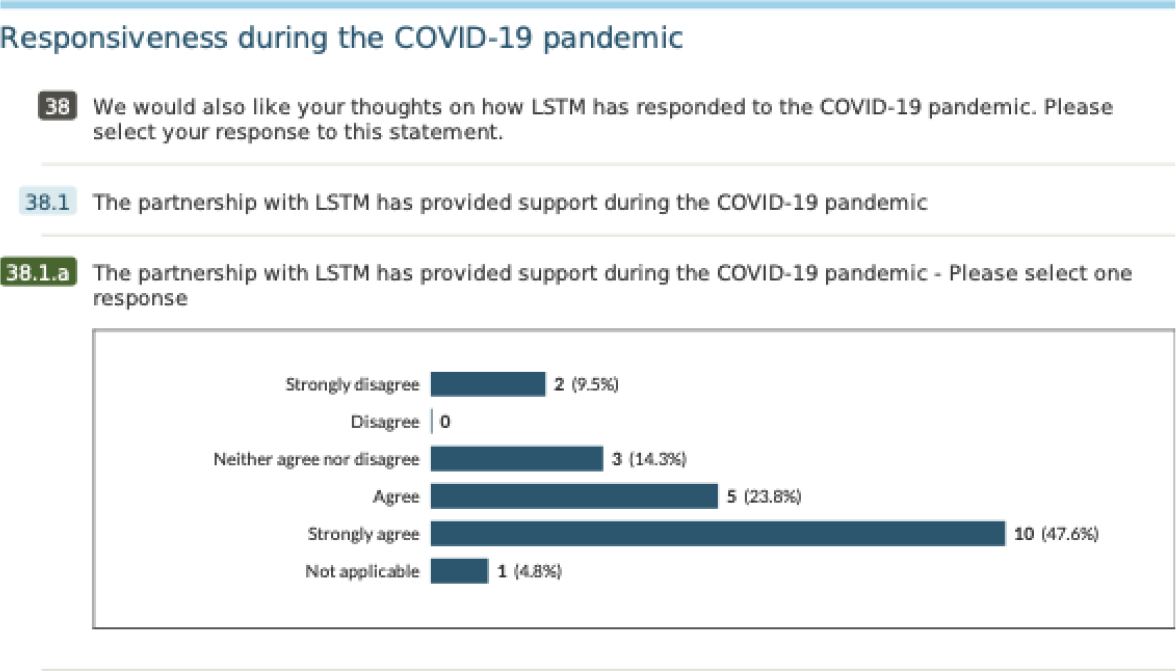

## Appendix 3: Topic Guide for Key Informant Interviews

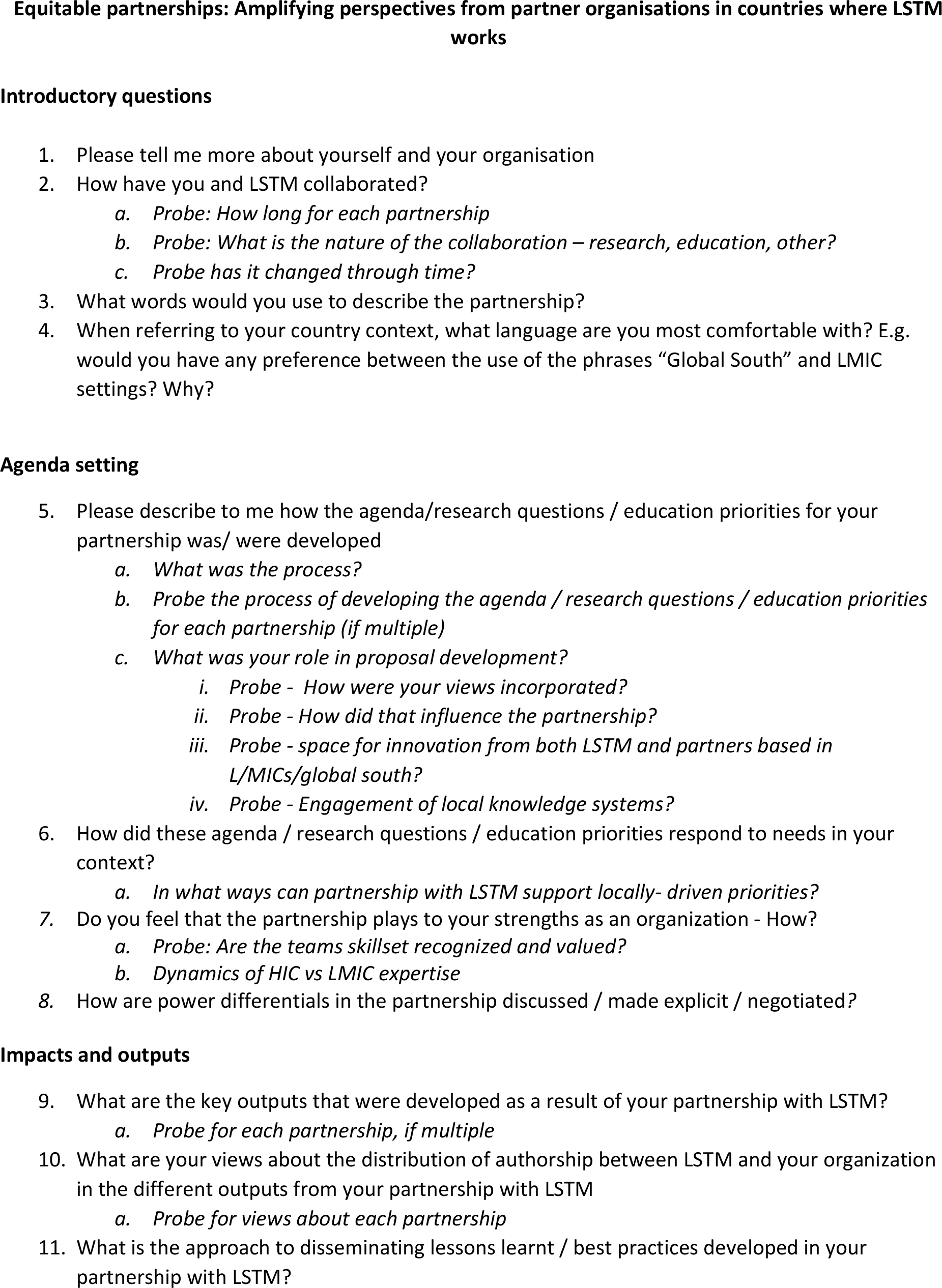
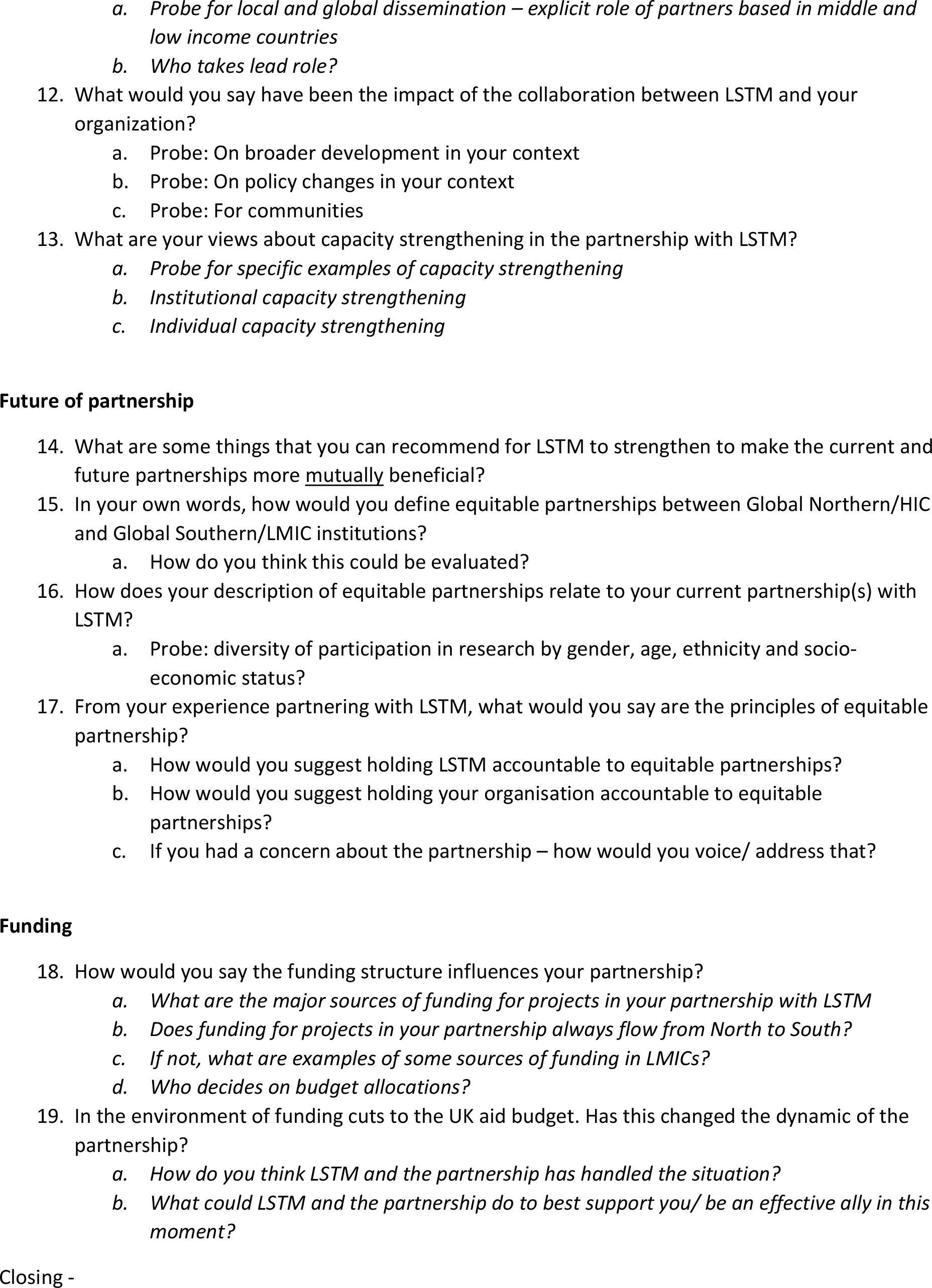
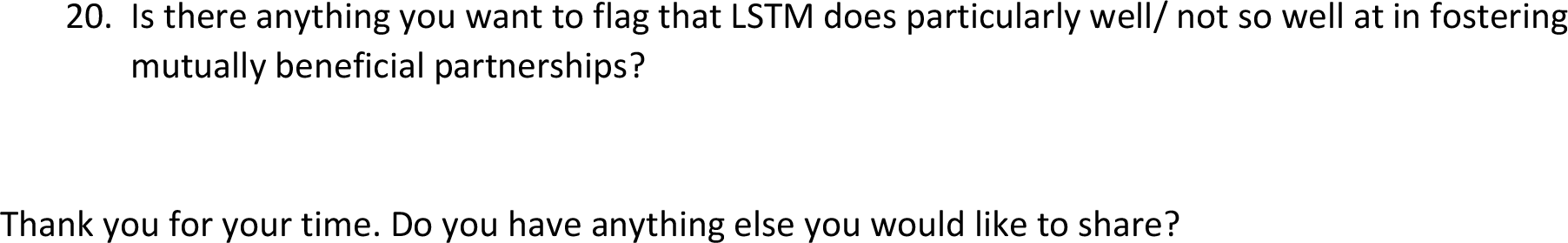

## Appendix 4: Consensus statement as per Morton et al. guidance on equitable authorship

1. How does the study address local research and policy priorities? Our study was specifically designed to support equity within transboundary partnerships. Supporting equitable partnerships is a key component of decolonising global health which is an issue of major priority in global health partnerships.
2. How were local researchers involved in study design? This was a global study designed by researchers in LSTM Liverpool (BS, ST, RS, SC) and LVCT health Kenya (RK and LO). LVCT Health and LSTM are longstanding partners and both are aligned in their goals to support equity in partnership. The study was designed to engage the perspectives of transboundary partners to co-develop principles for equitable partnerships in this area.
3. How has funding been used to support the local research team? The funding was used for pay for staff time in both LVCT Health and LSTM.
4. How are research staff who conducted data collection acknowledged? RS and RK conducted the data collection for the survey and KIIs, SC was also involved in data collection for the workshop. All research staff involved are authors on this paper.
5. Do all members of the research partnership have access to study data? All members of the partnership have access to data.
6. How was data used to develop analytical skills within the partnership? Most of the members of the research team are mid-career or senior researchers. SC is a research assistant and doctoral candidate and played a key role in workshop facilitation and analysis. She was supported by RS and RK. SC also presented on behalf of the research team at the Health Systems Global Conference in Bogota 2022.
7. How have research partners collaborated in interpreting study data? LVCT health colleagues (RK, LO) played a key role in analysing and interpreting the study data. In addition we:

- All participants were invited to a virtual workshop where the findings were presented for feedback followed by a process of co-creating principles. The principles were then also shared by email with all partners for any further additions or reflections.
- The study process, feedback and principles were also fed back to LSTM staff at a workshop and anonymous feedback was elicited through easy retro
- The study process, feedback and principles were also shared as part of a session on “Equitable Partnerships” held at the Health Systems Global conference in Bogota in Nov, 2022 and discussed in a world café format.
- Partners were also given opportunity to comment on the manuscript before submission
8. How were research partners supported to develop writing skills? We supported each other in the development of the outputs – including ethical protocols, PowerPoints and papers.
9. How will research products be shared to address local needs? We have opted for open access publication so that our research process and findings and principles are open for all to learn from and adapt as appropriate. These principles can be used by partners to hold LSTM to account in future collaborations.
10. How is the leadership, contribution and ownership of this work by LMIC researchers recognised within the authorship? The contribution of LVCT in shaping and conducting this research and output has been recognised with joint first (RK) and last authorship (LO) positions.
11. How have early career researchers across the partnership been included within the authorship team? We have included SC as a doctoral researcher within the authorship team, her critical role has been acknowledged as a corresponding author.
12. How has gender balance been addressed within the authorship? Four authors are women (RS, SC, ST, LO) and two men (RK, BS). We have women as both first author (RS) and last (LO/ST) author positions.
13. How has the project contributed to training of LMIC researchers? The LMIC researchers involved in the research are mid-senior in their career so this was not applicable.
14. How has the project contributed to improvements in local infrastructure? This project has not directly contributed to improvements in local infrastructure.
15. What safeguarding procedures were used to protect local study participants and researchers? We used the [LSTM safeguarding policy](https://www.lstmed.ac.uk/sites/default/files/LSTM%20Strategic%20Safeguarding%20Policy%20V3%2028.07.21.pdf) to guide this study; no safeguarding issues emerged during the study.

